# Cerebrovascular reactivity impairment in genetic frontotemporal dementia

**DOI:** 10.1101/2024.03.24.24304799

**Authors:** Ivana Kancheva, Arabella Bouzigues, Lucy L. Russell, Phoebe H. Foster, Eve Ferry-Bolder, John van Swieten, Lize Jiskoot, Harro Seelaar, Raquel Sanchez-Valle, Robert Laforce, Caroline Graff, Daniela Galimberti, Rik Vandenberghe, Alexandre de Mendonça, Pietro Tiraboschi, Isabel Santana, Alexander Gerhard, Johannes Levin, Sandro Sorbi, Markus Otto, Florence Pasquier, Simon Ducharme, Chris R. Butler, Isabelle Le Ber, Elizabeth Finger, Maria Carmela Tartaglia, Mario Masellis, Matthis Synofzik, Fermin Moreno, Barbara Borroni, Jonathan D. Rohrer, Louise van der Weerd, James B. Rowe, Kamen A. Tsvetanov, The GENFI consortium

## Abstract

**INTRODUCTION:** Cerebrovascular reactivity (CVR) is an indicator of cerebrovascular health and its signature in hereditary frontotemporal dementia (FTD) remains unknown. We investigated CVR in genetic FTD and its relationship to cognition.

**METHODS:** CVR differences were assessed between 284 pre-symptomatic and 124 symptomatic mutation carriers, and 265 non-carriers, using resting-state fluctuation amplitudes (RSFA) on component-based and voxel-level RSFA maps. Associations and interactions between RSFA, age, genetic status, and cognition were examined using generalised linear models.

**RESULTS:** Compared to non-carriers, mutation carriers exhibited greater RSFA reductions, predominantly in frontal cortex. These reductions increased with age. The RSFA in these regions correlated with cognitive function in symptomatic and, to a lesser extent, pre-symptomatic individuals, independent of disease stage.

**DISCUSSION:** CVR impairment in genetic FTD predominantly affects frontal cortical areas, and its preservation may yield cognitive benefits for at-risk individuals. Cerebrovascular health may be a potential target for biomarker identification and disease-modifying efforts.

## 1. Background

Frontotemporal dementia (FTD) encompasses a clinically heterogeneous group of neurodegenerative diseases [1]. About a third of FTD cases present an autosomal dominant family history, commonly caused by mutations in three genes: chromosome 9 open reading frame 72 (*C9orf72*), progranulin (*GRN*), and microtubule-associated protein tau (*MAPT*) [2]. The study of prodromal FTD has identified neuropathological changes and biomarker abnormalities decades before disease onset, including brain atrophy, reduced white matter (WM) integrity, and disrupted functional connectivity, predominantly affecting the fronto-temporo-parietal regions [3].

In addition to the tau and TDP-43-associated molecular pathologies, and secondary inflammation, the pathophysiology of FTD involves cerebrovascular dysregulation [4]. It is characterised by impairments in the brain’s neurovascular unit (NVU) and blood-brain barrier (BBB), with damaged endothelial cells, dysfunctional pericytes, and adjacent reactive microglia, in people carrying FTD-related mutations [5, 6]. Furthermore, reductions in cerebral blood flow (CBF) are found in both sporadic and genetic FTD, especially in frontal cortex [7, 8]. The changes in cerebral blood flow correlate with impaired performance on neuropsychological tests [9]. Combined with evidence of small-vessel pathology in autopsy-confirmed cases with frontotemporal lobar degeneration (FTLD) [10], these findings suggest a synergistic contribution of neurodegeneration and cerebrovascular impairment to the pathophysiology of FTD [4].

An important aspect of cerebrovascular function is cerebrovascular reactivity (CVR). CVR denotes the capacity of cerebral blood vessels to constrict or dilate in response to physiological modulators, such as carbon dioxide concentration [11]. CVR regulates regional blood flow via pH-dependent modulation of vascular smooth muscle tone [12–14]. It is compromised by ageing [15], impaired endothelial function [16], and hypertension [17]. The blood oxygenation-level dependent (BOLD) contrast reveals CVR alterations in Alzheimer’s disease (AD) and its prodrome [18, 19], leading to the hypothesis of comparable FTD-related changes in CVR.

In this study, we investigated CVR in pre-symptomatic and symptomatic genetic FTD. We used existing resting-state functional magnetic resonance imaging (rs-fMRI) data that are based on exploiting naturally occurring fluctuations in carbon dioxide, induced by variations in the cardiac and respiratory cycles, which moderate the BOLD signal [20, 21]. Resting-state fluctuation amplitudes (RSFA) of the BOLD signal is a safe, scalable, and robust alternative to the standard MRI approaches [22–24]. It is especially suitable for large-scale applications with frail subjects, as it does not require hypercapnic gas inhalation, breath-holding, or vasodilatory drugs [24–25], for a review, see Tsvetanov et al. (2021) [26]. RSFA has already been used to assess differences in cerebrovascular and cardiovascular function associated with ageing [27–29], cerebrovascular disorders [30], stroke [31], AD [32], as well as other acute conditions that might heighten the risk of dementia [33].

The principal aim was to determine the CVR signature of pre-symptomatic and symptomatic genetic FTD. A corollary was to assess CVR correlations with age and clinical status. We predicted reductions in RSFA in at-risk mutation carriers compared to mutation-negative family members; and that these differences would increase with disease progression and relate to impaired cognitive performance.

## 2. Methods

### 2.1. Participants

Data were drawn from the fifth data freeze of the Genetic Frontotemporal Dementia Initiative (GENFI, www.genfi.org), which included 31 research sites across Europe and Canada. The study was approved by the institutional review boards at each site and written informed consent was provided by participants. A total of 680 subjects were recruited between January 30, 2012, and May 28, 2019, from families with a confirmed pathogenic genetic mutation in *C9orf72*, *GRN*, or *MAPT*. They were either (i) symptomatic mutation carriers, (ii) first-degree relatives of mutation carriers who were carrying a mutation, but did not exhibit any symptoms (that is, pre-symptomatic), or (iii) mutation-negative family members who served as a control group, termed non-carriers. Subjects were classified as symptomatic if their clinician judged the presence of symptoms consistent with the diagnosis of a progressive in nature degenerative disorder. Seven datasets were excluded due to motion-related or other imaging artifacts (three symptomatic subjects with *C9orf72* mutations; three pre-symptomatic *GRN* carriers, and one mutation-negative individual from a family with a *GRN* mutation). This resulted in 673 usable fMRI scans from 124 symptomatic mutation carriers (61 *C9orf72*, 40 *GRN*, 23 *MAPT*), 284 pre-symptomatic mutation carriers (107 *C9orf72*, 123 *GRN*, 54 *MAPT*), and 265 mutation-negative controls.

### 2.2. Neurocognitive Assessment and Indices of Cognitive Function

All participants underwent the standardised GENFI clinical evaluation consisting of family and medical history, functional status, and physical examination in corroboration with collateral history from a close contact. Subjects also completed a neuropsychological battery, which included behavioural measures of cognitive function from the Uniform Data Set [34]. From this test battery, we used scores related to executive function (Digit Span Forwards and Backwards from the Wechsler Memory Scale-Revised; Parts A and B of the Trail Making Test; a Digit Symbol Task) and language (the short version of the Boston Naming Test; Category Fluency (animals and combined)), as well as the Wechsler Abbreviated Scale of Intelligence Block Design Task. More details on the recruitment procedure and clinical assessment protocol can be found in Rohrer et al. (2015) [35].

As a proxy of cognitive function for subsequent statistical analysis, we used Principal Component Analysis (PCA) to derive a latent variable from a set of cognitive performance assessments. This enabled us to obtain a composite summary score characterising the complexity of cognition whilst minimising the statistical problem of multiple comparisons when investigating associations between genetic status, RSFA, and cognitive function. Thus, we conducted PCA on subjects’ performance measures from the Digit Span Forwards and Backwards task, Parts A and B of the Trail Making Test, the Digit Symbol Task, Boston Naming Test, Category Fluency (animals and combined), and Block Design Task to reduce the dimensionality of cognitive function into one latent variable summarising the largest portion of shared variance as the first principal component (PC 1). In cases of missing values for some of the metrics, multivariate Markov Chain Monte Carlo imputation was performed using the default settings of the multivariate imputation by chained equations (MICE) in R [36].

### 2.3. Image Acquisition and Pre-processing

A three-dimensional (3D) structural MRI was obtained for each participant using a T1-weighted Magnetic Prepared Rapid Gradient-Echo (MPRAGE) sequence on 3T scanners available from various vendors. The scanning protocols at each GENFI site were optimised to accommodate different manufacturers and field strengths [35]. The following acquisition parameters were used: median isotropic resolution of 1 mm; repetition time (TR) of 2000 ms (6.6 to 2400 ms); echo time (TE) of 2.9 ms (2.8 to 4.6 ms); inversion time (TI) of 8 ms (8 to 9 ms); field of view (FOV) of 256 × 256 × 208 mm, with a minimum scanning time of at least 283 s (283 to 462 s).

The T1-weighted images were analysed using FSL pipelines [37, 38] and modules, which called relevant functions from Statistical Parametric Mapping (SPM12, Wellcome 112 Department of Imaging Neuroscience, London, UK; www.fil.ion.ucl.ac.uk/spm) [39]. Native-space segmentation of grey matter (GM), WM, and cerebrospinal fluid (CSF) tissue classes and warps for normalisation to the Montreal Neurological Institute (MNI) template space were estimated using FSL.

For resting-state fMRI measurements, Echo-planar imaging (EPI) data were obtained with at least six minutes of scanning. Analogous imaging sequences were developed by the GENFI Imaging Core team and used at each GENFI study site to account for different scanner models and field strengths. EPI data were acquired over a minimum of 308 s (median 500 s) and had a median TR of 2500 ms (2200 to 2500 ms); TE of 30 ms; flip angle of 80 ms (80 to 85 ms); in-plane resolution of 2.72 × 2.72 mm (2.72-3.50 × 2.72-3.25 mm); slice thickness of 3.5 mm (2.72 to 3.5 mm). Participants were instructed to lie still with their eyes closed. The initial six volumes were discarded to enable T1 equilibration. To quantify the total motion for each participant, the root mean square volume-to-volume displacement was computed using the approach of Jenkinson et al. (2002) [40].

The pre-processing was carried out using SPM12 running under MATLAB R2021b (MathWorks, https://uk.mathworks.com/). The pre-processing steps comprised (i) spatial realignment to correct for head motion and movement by distortion interactions, (ii) slice-time correction to the middle slice, (iii) co-registration of the EPI to the participants’ T1 anatomical scans. The normalisation parameters from the T1 image processing were then applied to warp the functional images to MNI space. After that, the spatially normalised images were smoothed with a Gaussian kernel of Full Width at Half Maximum (FWHM) of 8 mm to meet the lattice assumption of random field theory and account for residual inter-participant structural variability.

Further processing procedures of the resting-state time-series for estimation of RSFA involved the application of data-driven Independent Component Analysis (ICA) of single-subject time-series denoising, with noise components selected and removed automatically using *a priori* heuristics from the ICA-based Automatic Removal of Motion Artifacts (AROMA) toolbox [41]. A general linear model (GLM) of the time-course at each voxel was computed to further diminish any residual effects of noise confounds [42]. This included linear and quadratic detrending of the fMRI signal, covarying out the motion parameters, WM, and CSF signals, as well as their squares and first derivative [43], and a band-pass filter (0.0078–0.01 Hz). Signals from WM and CSF were estimated for each volume from the mean value of WM and CSF masks derived by thresholding SPM’s corresponding tissue probability maps at 0.75. The normalised variance (i.e., the temporal standard deviation (SD) of the EPI signal amplitudes over the mean signal intensity) of these filtered time-series was calculated on a voxel-wise basis to define RSFA.

### 2.4. Indices of Cerebrovascular Function using RSFA

#### 2.4.1. Component-based Analysis

To implement ICA, the pre-processed RSFA maps were decomposed into a set of spatially independent sources using the Source-Based Morphometry toolbox [44] in the Group ICA for fMRI Toolbox (GIFT; http://mialab.mrn.org/software/gift). Briefly, the fastICA algorithm was applied after the optimal number of sources explaining the variance in the data was identified by PCA with Minimum Description Length (MDL) criterion [45–47]. By combining PCA and ICA, one can decompose an n-by-m matrix of subjects-by-voxels into a source matrix that maps independent components (ICs) to voxels (here referred to as ‘IC maps’), and a mixing matrix that maps ICs to participants. The mixing matrix indicates the degree to which an individual expresses a defined IC, known as the subject scores in the mixing matrix. These scores were scaled to standardised values (z-scores) prior to between-group analyses. The algorithmic and statistical reliability of the extracted components was confirmed with 128 ICASSO (tool for investigating the reliability of ICA estimates by clustering and visualisation) iterations [48]. Components showing high reliability across multiple ICASSO iterations and comprising GM areas were deemed relevant and used in subsequent analyses. This decision was based on the understanding that RSFA alterations within GM areas are indicative of cerebrovascular reactivity (CVR) [29] and are shown to be sensitive to cognitive function [49].

#### 2.4.2. Voxel-based Univariate Analysis

To understand the spatial distribution of CVR effects further, we performed a sensitivity voxel-wise analysis on the RSFA maps using SPM12 in MATLAB. Clusters where between-group RSFA differences were observed after correction for multiple testing were used to define regions of interest (ROIs) for visualisation of effects across subjects.

### 2.5. Statistical Analysis

#### 2.5.1. Descriptive Statistics

Demographic characteristics were compared with SPSS (IBM Corp. Released 2021. IBM SPSS Statistics for Windows, Version 29.0. Armonk, NY: IBM Corp). Due to unequal sample sizes and variances between groups, Welch’s ANOVA with Games-Howell post hoc tests were employed for continuous data. Chi-square tests were used for categorical variables. The significance level was defined as two-tailed, and the threshold was set at *p* = 0.05 for all statistical procedures. In keeping with other GENFI reports, years to expected onset (EYO) was defined as the difference between age at assessment and mean age at onset within the family [35]. EYO is only provided for completeness and should be interpreted with caution, noting that age of symptom onset cannot be reliably predicted based on family history in *GRN* and *C9orf72* mutation carriers [50].

#### 2.5.2. FTD-related Effects on Cerebrovascular Indices using RSFA

Figure 1 gives a schematic overview of the processing pipeline and analytic approach used in the study. RSFA differences between symptomatic and pre-symptomatic mutation carriers (all genetic mutations combined) and non-carriers were examined on component-based estimates of RSFA using multiple linear regression (MLR) with a robust fitting algorithm (MATLAB function *fitlm.m*). In these models, the IC subject scores for each component (termed RSFA_IC□_, where *n* denotes the corresponding number of the selected component) were entered as a dependent variable, with age, sex, and handedness as covariates of no interest. Although scanning protocols within the cohort were designed to maximise comparability across GENFI scanners and sites [35], distinct scanning platforms can introduce systematic differences, potentially confounding true effects of interest [51]. Thus, scanning site was also inserted as a covariate of no interest.

**Figure 1.**
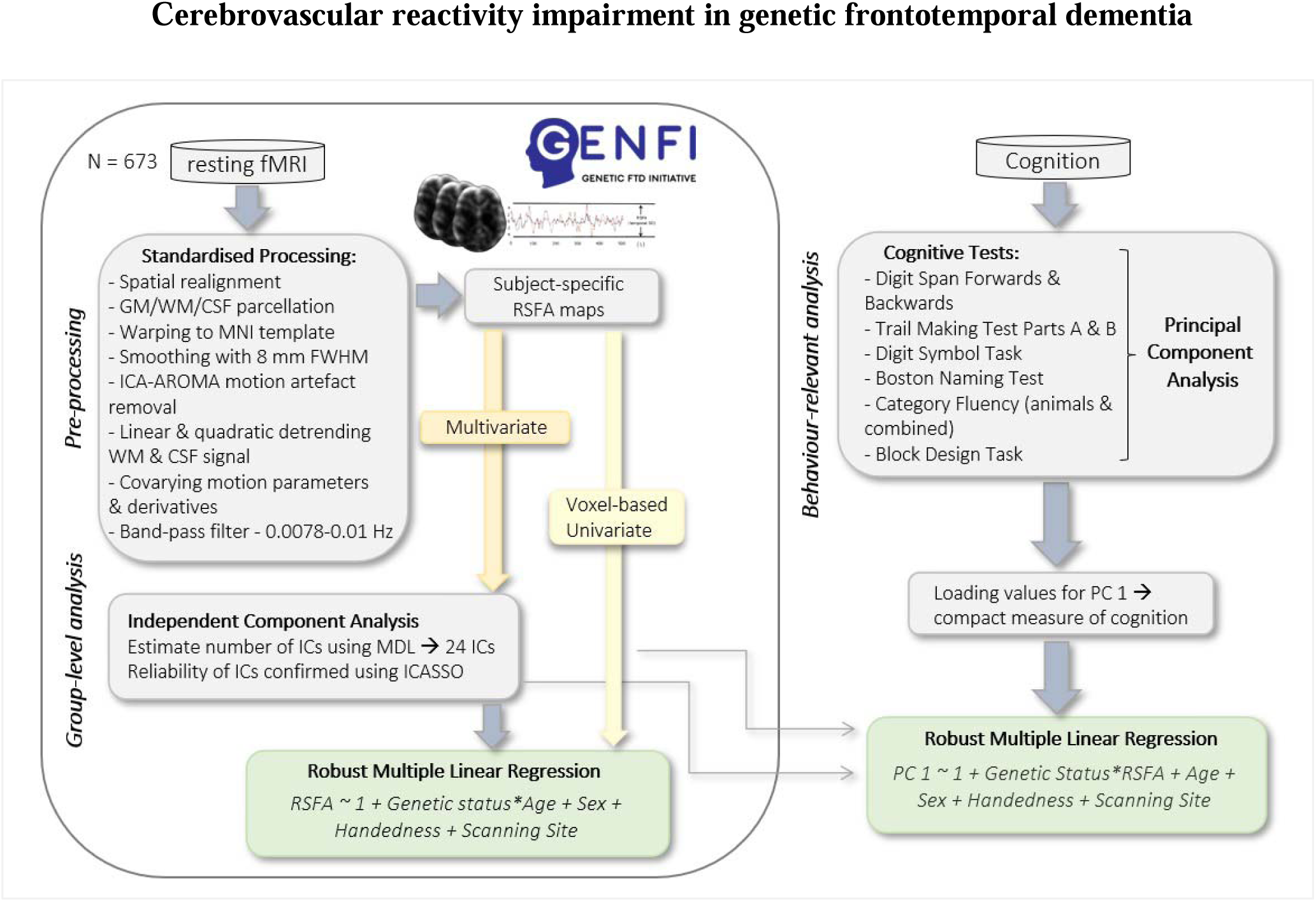
Schematic representation of the pre-processing pipeline and analytic approach used in the study.

We also tested the moderating effect of age on the case-control differences to explore disease progression-related effects across genetic status groups, i.e., whether the age effect in pre-symptomatic carriers would be stronger than the ‘normal’ age effect in non-carriers, due to the development of latent, pre-symptomatic pathology. This enabled us to assess the variance explained by genetic status beyond that accounted for by age and other covariates in the MLR.

All models’ formulas were specified by Wilkinson’s notation, e.g., *‘RSFA_IC_ ∼ 1 + Genetic status*Age + Sex + Handedness + Scanning Site’*, providing a flexible way to test for main effects of predictors of interest (i.e., genetic status and age) and their interaction (genetic status*age), whilst adjusting the models for confounders of no interest (sex, handedness, scanning site). To account for issues related to multiple testing, the overall model fit was corrected using the Benjami-Hochberg procedure to control the false discovery rate (FDR) at the 0.05 level. To better understand the nature of the observed effects in the models surviving multiple comparisons, post hoc tests were performed across sub-groups of interest (e.g., comparing non-carriers to symptomatic carriers, non-carriers to pre-symptomatic carriers, and pre-symptomatic carriers to symptomatic carriers).

A sensitivity analysis was also conducted on the voxel-wise RSFA maps to further explore the spatial distribution of CVR effects within the same statistical model (*‘RSFA_Voxel_ ∼ 1 + Genetic status*Age + Sex + Handedness + Scanning Site’)*. This included the following comparisons: (i) non-carriers versus symptomatic carriers, (ii) non-carriers versus pre-symptomatic carriers, and (iii) pre-symptomatic carriers versus symptomatic carriers. The primary cluster-forming threshold was set at *p* = 0.05. To correct for the multiple comparisons problem inherent in mass univariate statistical analysis, we controlled for the voxel-level FDR at *p* < 0.05. For transparency, in cases where results did not reach statistical significance at FDR-level, the patterns are reported at an uncorrected level of *p* < 0.01 with a minimum cluster size of 10 voxels. To visualise the nature of the observed effects and any further between-group RSFA differences, ROIs were defined by selecting the voxels in an 8-mm sphere at the peak of significant clusters from the MLR analysis. The average value across voxels in each ROI was used to illustrate the relationship between RSFA and age for different genetic status groups. The voxel-based level regions were labelled according to their overlap with the Johns Hopkins University (JHU) atlas [52].

#### 2.5.3. Behavioural Relevance of Cerebrovascular Impairment

A secondary objective of this study was to evaluate the behavioural relevance of the RSFA changes observed in the previous analyses for subjects’ cognitive function. To assess differences in cognitive performance scores between genetic status groups, we first carried out a Kruskal-Wallis test, followed by Mann-Whitney post hoc tests. We subsequently ran a further series of regression models where cognitive function, as represented by subjects’ scores for PC 1 from the PCA analysis, was the dependent variable. Independent variables included the RSFA_IC_ for each neurocognitively meaningful component (see Methods, 2.4. Indices of Cerebrovascular Function using RSFA, sub-section 2.4.1. Component-based Analysis), and its interaction with genetic status, to test whether the relationship between cognitive performance and RSFA levels would vary across genetic status groups. Covariates of no interest were age, sex, handedness, and scanning site. Models’ formulas, as specified by Wilkinson’s notation, took the form: *‘Cognition_PC1_ ∼ 1 + Genetic status*RSFA_IC/Voxel_ + Age + Sex + Handedness + Scanning Site’*. FDR correction was applied (FDR < 0.05) and post hoc tests across sub-groups of interest were conducted in cases where main effects were found (Figure 1).

## 3. Results

### 3.1. Demographics

Demographic characteristics of the sample and descriptive statistics are provided in Table 1. Mutation-negative family members and pre-symptomatic mutation carriers were younger than symptomatic carriers (mean difference between non-carriers and symptomatic carriers was 16.68 years (*p* < 0.001) and 18.71 years between pre-symptomatic carriers and symptomatic carriers, respectively, (*p* < 0.001)). The pre-symptomatic carriers were age-matched to non-carriers (*p* = 0.132). There was a higher proportion of females, relative to males, in asymptomatic individuals (non-carriers and pre-symptomatic carriers) compared to symptomatic carriers, and these two groups had also spent more years in education. No significant differences were observed between the non-carriers and pre-symptomatic carriers for any of the remaining demographic variables.

**Table 1.** Demographic information of participants included in the analysis, grouped by genetic status as non-carriers, pre-symptomatic carriers, and symptomatic carriers

| Demographics | Sample | NC | PSC | SC | Group comparison,<br>*P* value* |  |  |  |
| --- | --- | --- | --- | --- | --- | --- | --- | --- |
|  |  |  |  |  | *Sample* | *NC vs SC* | *PSC vs SC* | *NC vs PSC* |
| Total N | 673 | 265 (39.38) | 284 (42.2) | 124 (18.42) |  |  |  |  |
| Family mutation |  |  |  |  | .126 |  |  |  |
| *C9orf72* | 264 (39.23) |  | 107 | 61 |  |  |  |  |
| *GRN* | 276 (41.01) |  | 123 | 40 |  |  |  |  |
| *MAPT* | 133 (19.76) |  | 54 | 23 |  |  |  |  |
| Age (years) | 48.17±13.43 | 45.95±13.09 | 43.93±11.4 | 62.64±7.43 | **<.001** | **<.001** | **<.001** | .132 |
| Sex ratio f:m | 371:302 | 153:112 | 165:119 | 53:71 | **.009** | **.006** | **.004** | .931 |
| Estimated years from onset | -10.62±13.40 | -13.21±13.47 | -14.30±11.63 | 3.32±6.24 | **<.001** | **<.001** | **<.001** | .569 |
| Education (years) | 14.18±3.45 | 14.51±3.35 | 14.50±3.36 | 12.72±3.53 | **<.001** | **<.001** | **<.001** | .998 |

### 3.2. Regional Differences in RSFA based on Independent Component Analysis

Applying ICA to the RSFA data yielded 24 components based on the MDL criterion. The spatial patterns indicated signal origins within GM regions, as well as origins associated with vascular aetiology, CSF, or other non-physiological factors (Appendix A, Figure A.1). A total of 20 components were excluded from further analysis; these included non-GM components proximal to vascular and CSF territories or components exhibiting characteristics of physiological noise signals (Appendix A, Table A.1). Additionally, GM components that did not survive correction for multiple comparisons were also classified as irrelevant. The overall model fit of four GM components remained significant after FDR correction (Figure 2). These components included strong contributions of voxels within the posterior cingulate cortex (PCC)/precuneus (IC 4), posterior association and parieto-occipital association areas, more pronounced on the right side (IC 17), right and left lateral prefrontal cortex (IC 21 and IC 23, respectively). A tendency of FTD-dependent decrease in RSFA was found in symptomatic and pre-symptomatic carriers, compared to non-carriers, for all components, but only reached statistical significance for component IC 21. Post hoc tests revealed that this effect was driven by differences between the non-carriers and symptomatic mutation carriers, as well as between the pre-symptomatic and symptomatic carriers. In addition, in analyses across the entire sample, a significant main effect of genetic status x age interaction was shown for components IC 17, IC 21, and IC 23, whereby symptomatic carriers showed the most pronounced age-related RSFA reductions, followed by the presymtompatic carriers, and then mutation-negative individuals. This suggests a greater age-related RSFA decline in at-risk or affected mutation carriers relative to non-carriers, which likely further exacerbates downstream effects of the disease over time, as indicated by the steeper negative slopes of the regression lines observed in these groups. Spatial maps of these components, accompanied by scatter plots showing IC subject score values in relation to age and genetic status group, are presented in Figure 2. Table 2 summarises the numerical output of the MLR models conducted on the RSFA-IC subject scores for these GM components across the entire sample. The output of post hoc tests across sub-groups of interest for this analysis is provided in Appendix A, Table A.2.

**Figure 2.**
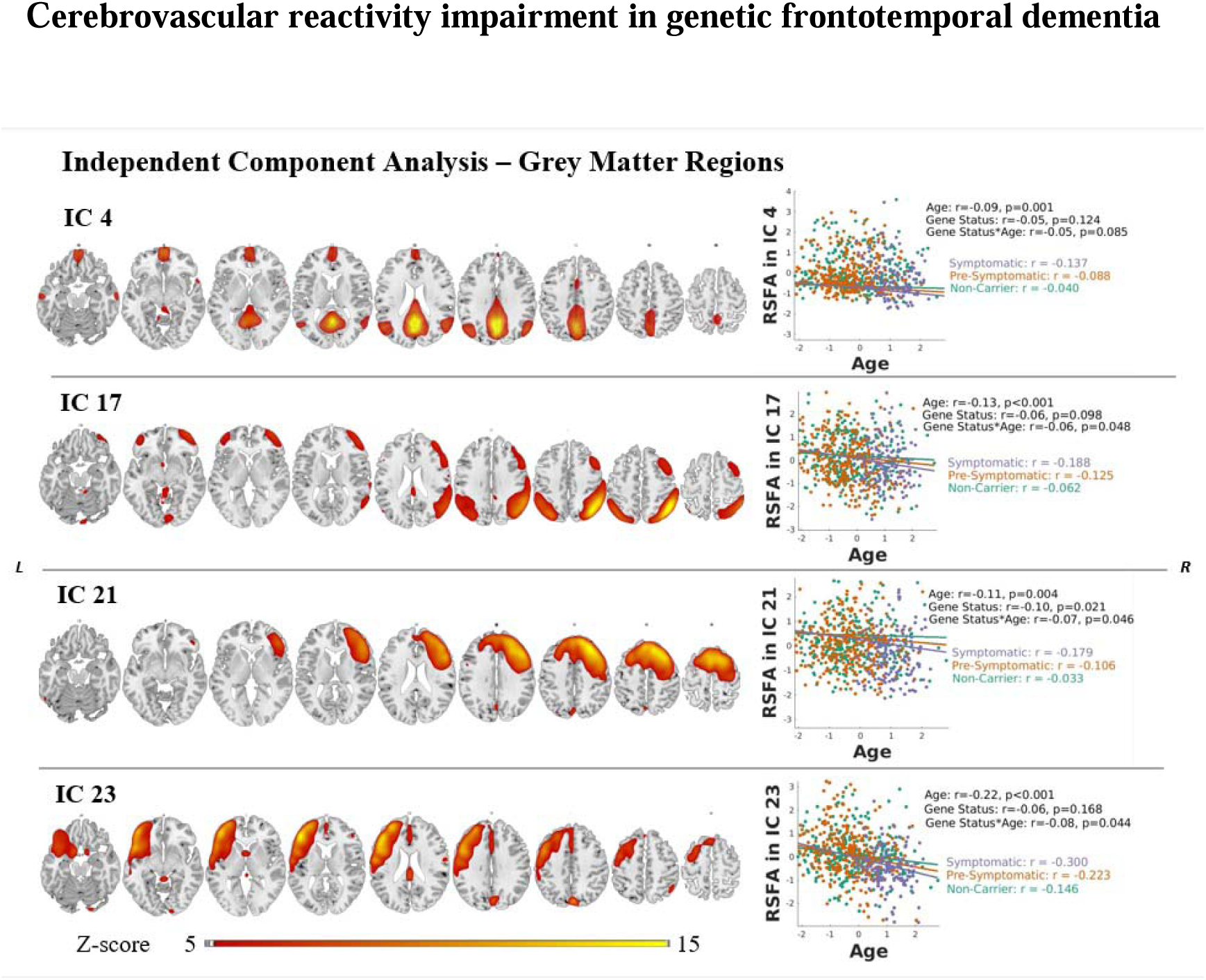
Spatial distribution of 4 independent components (ICs) within neurocognitively meaningful areas (i.e., GM regions) based on ICA on RSFA maps across subjects where differences in IC loading values are found in association with genetic status, age, and genetic status x age interaction. Robust general linear model regression lines for each IC are presented in scatter plots with respective r values on the right side of each IC map. P values are FDR-corrected at the 0.05 level across the whole sample. Group-level spatial maps are overlaid onto the Colin-27 (ch2.nii) structural template of the MNI brain, where intensity values correspond to z-values.

**Table 2.**
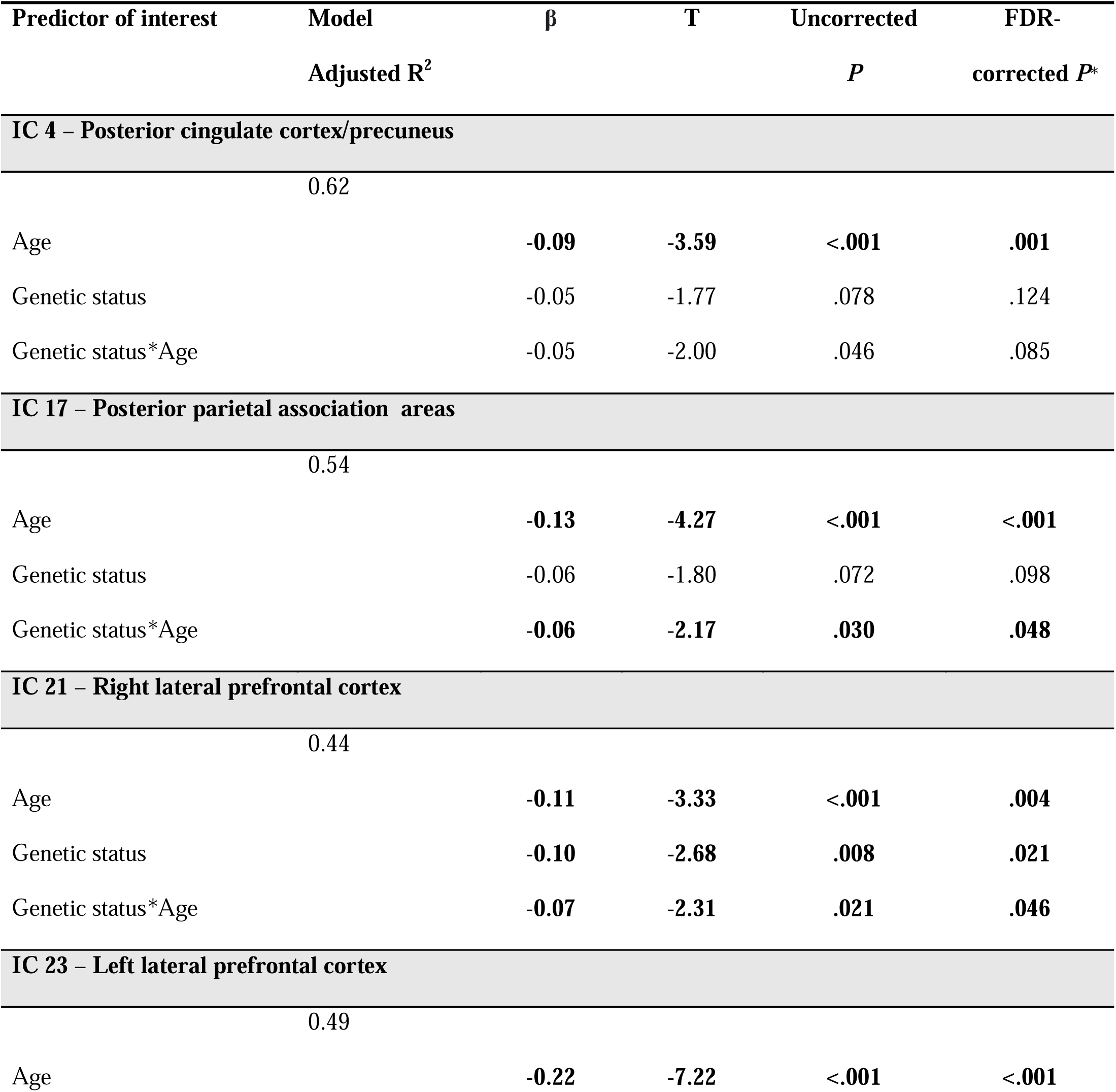
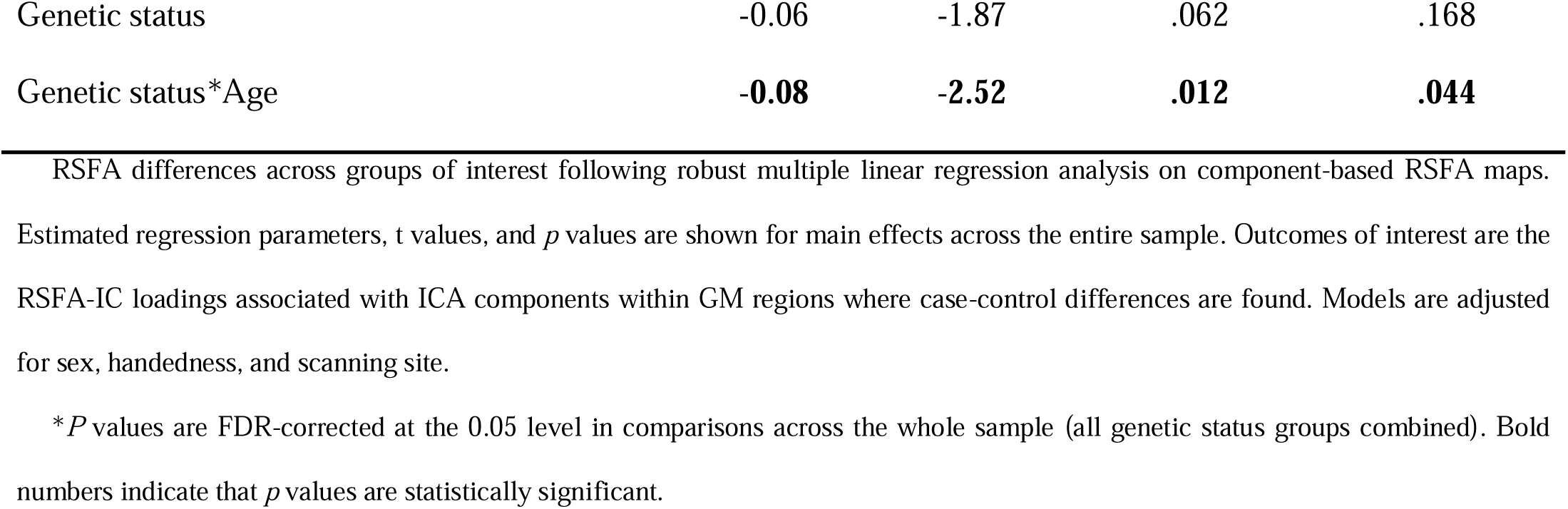
Multiple regression analysis results of independent component (IC) subject loadings from independent component analysis (model *‘RSFA_IC_ ∼ 1 + Genetic status*Age + Sex + Handedness + Scanning Site’*)

ICA also revealed components with spatial distribution originating from large blood vessels and CSF (Appendix A, Figure A.1). For example, components 2 and 3 reflected signals from the fluid-filled ventricles and cerebral aqueduct. Component 11 indicated signals originating close to sites of venous drainage, including superior and inferior sagittal sinus, and transverse sinuses. Other vascular components comprised territories of major blood vessels, including the Circle of Willis, internal carotid artery, anterior cerebral artery, and middle cerebral artery. These vascular and CSF components tended to display higher subject scores in older (symptomatic) individuals, reflecting differences in vascular health and other physiological factors [26, 28, 29].

### 3.3. Spatial Distribution and Voxel-wise Univariate Differences in RSFA

Overall, voxel-based analysis results were consistent with component-based analysis, particularly in frontal cortical regions (**Error! Reference source not found.** 3). Group-level analysis across all genetic groups revealed a consistent pattern of RSFA decreases in frontal midline areas, cuneus, precuneus, and cerebellum. Moreover, a comparable tendency emerged in relation to disease progression informed by the interaction between genetic status and age. Specifically, the inverse relationship between RSFA and age was stronger across the spectrum from non-carriers to pre-symptomatic carriers to symptomatic carriers. The spatial distribution of voxel-based RSFA effects, including four representative ROIs where the strongest effects were demonstrated, is illustrated in Figure 3. The output from the MLR models performed on the RSFA-ROI estimates across the entire sample is provided in Table 3. The results of post hoc tests across sub-groups of interest conducted subsequently are provided in Appendix A, Table A.3.

**Figure 3.**
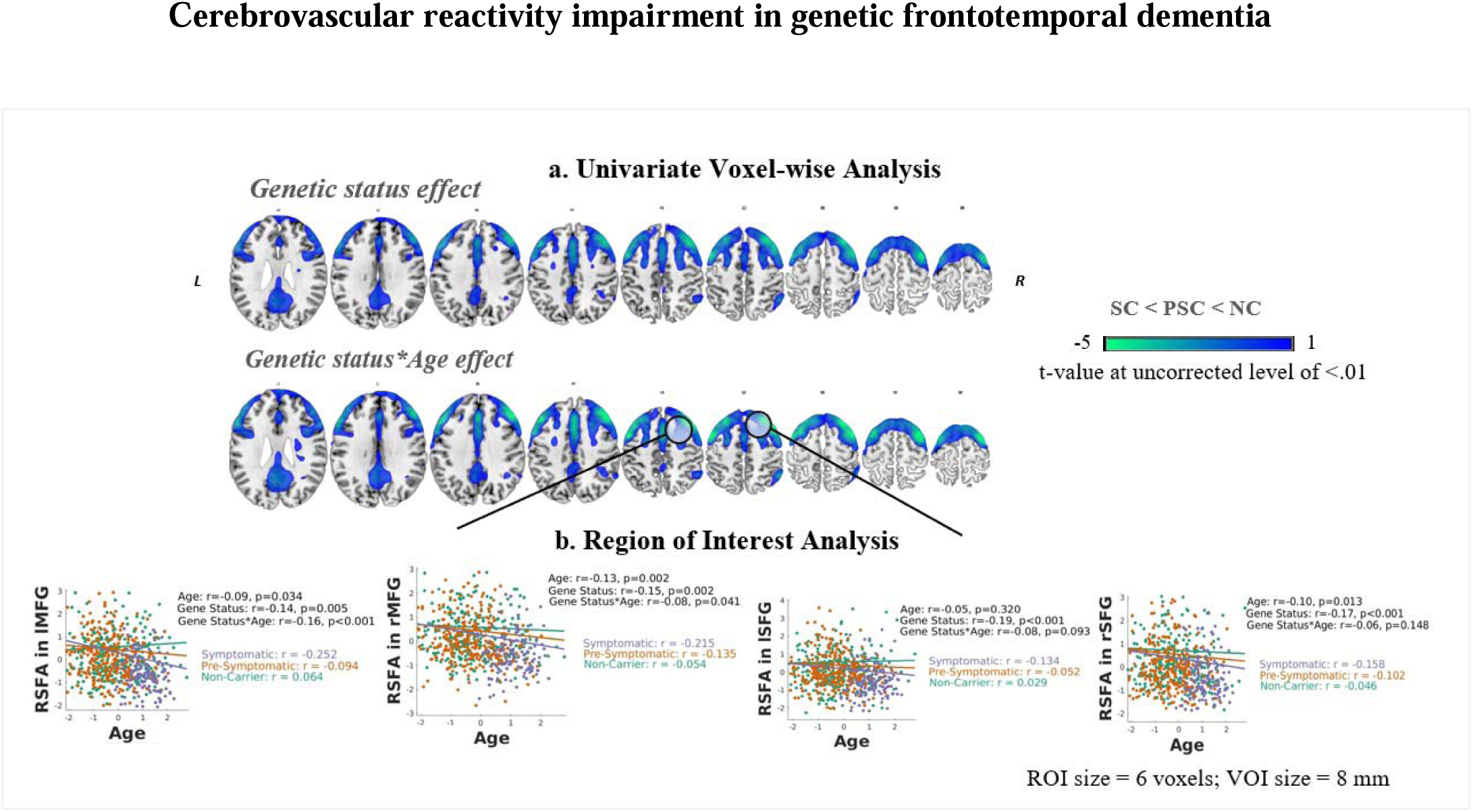
Panel A. Regional distribution of RSFA effects based on voxel-wise univariate analysis. Cold colours denote RSFA decreases as a function of genetic status and their interaction with age. Statistical parametric maps are displayed at an uncorrected level of p < 0.01 to better visualise regional CVR patterns. Images are overlaid onto the Colin-27 (ch2.nii) structural template of the MNI brain. Panel B. Differences in RSFA in association with genetic status, age, and genetic status x age interaction across groups of interest in several representative ROIs based on voxel-wise univariate analysis. Robust general linear model regression lines for each ROI are presented in scatter plots with respective r values on the right side of each ROI map. P values are FDR-corrected at the 0.05 level across the whole sample. NC, non-carrier; PSC, pre-symptomatic mutation carrier; SC, symptomatic mutation carrier. MFG, middle frontal gyrus; SFG, superior frontal gyrus. ROI, region of interest; VOI, volume of interest.

**Table 3.**
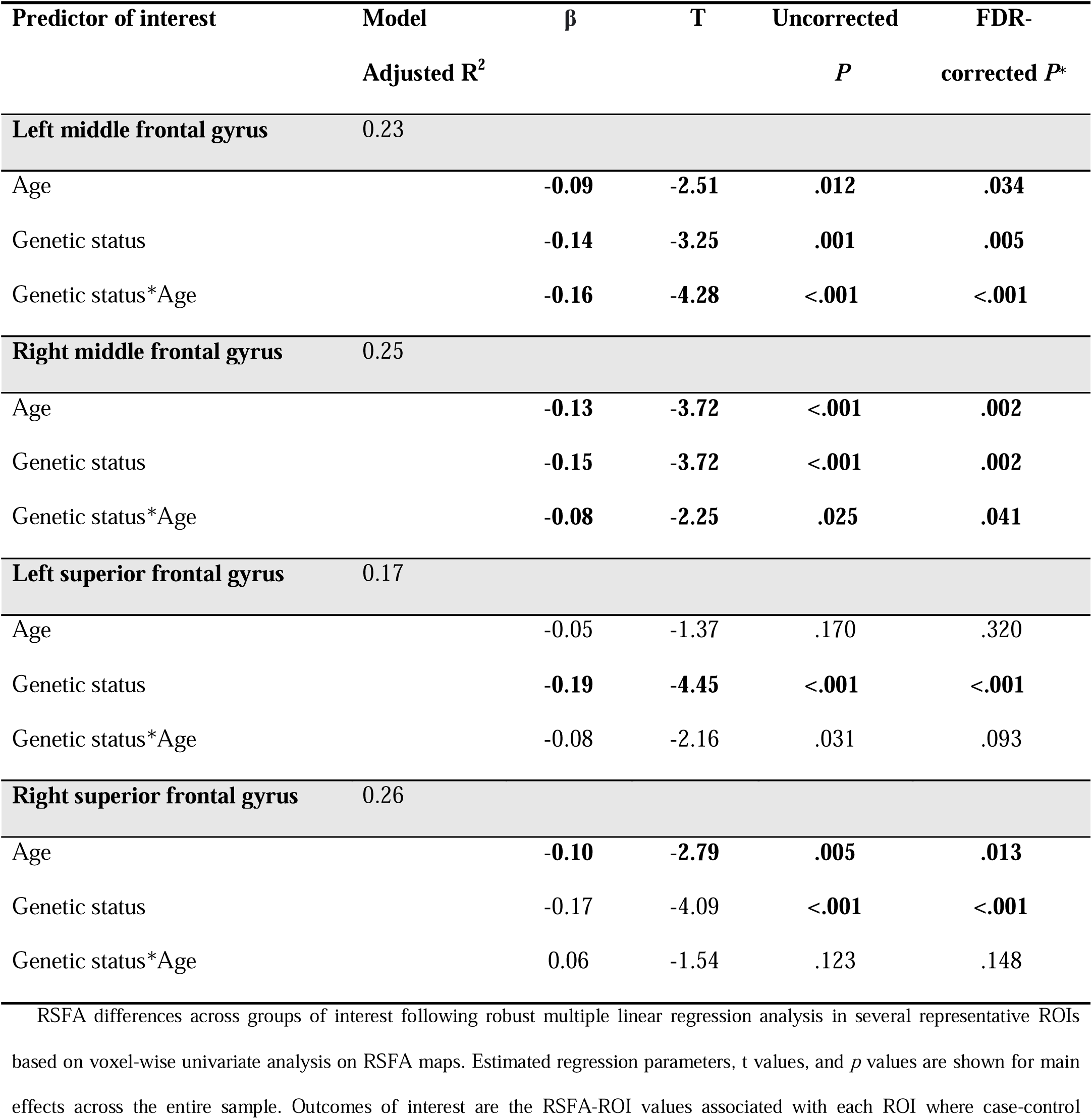
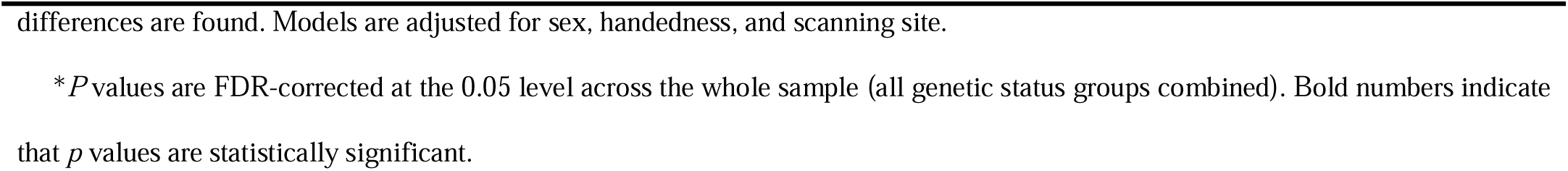
Multiple regression analysis results following voxel-based region of interest analysis (model *‘RSFA_Voxel_ ∼ 1 + Genetic Status*Age + Sex + Handedness + Scanning Site’*)

Regarding genetic status effects on voxel-wise RSFA, there were several clusters where symptomatic carriers exhibited significant reductions, compared to non-carriers, including bilateral middle frontal gyrus (MFG), right superior frontal gyrus (SFG), right superior temporal gyrus (STG), and bilateral PCC. Symptomatic carriers also displayed greater age-related RSFA decline in the same areas, as well as in the left SFG, left dorsal anterior cingulate cortex (ACC), and right insula. Similar clusters displayed RSFA decreases when symptomatic carriers were compared to pre-symptomatic counterparts. In contrast, the differences between pre-symptomatic carriers and non-carriers did not reach statistical significance at FDR-levels. However, pre-symptomatic carriers showed a tendency for reduced RSFA in posterior parietal cortex and more pronounced age-related decline in RSFA over the parietal and frontal cortex compared to non-carriers, similar to the trend observed in the symptomatic group. A detailed description of the anatomical localisation of the voxel-based analysis derived clusters where RSFA differences were noted can be consulted in Appendix A, Table A.4.

Finally, to evaluate differences in RSFA values across different gene mutations, we compared RSFA-IC loadings and RSFA-ROI estimates in mutation carriers stratified by gene mutation using MLR. No between-group differences were detected based on mutated gene in any of the ICs or ROIs where differences between asymptomatic and symptomatic carriers were encountered in the previous analyses (data are shown in Appendix A, Table A.5).

### 3.4. Relationship between RSFA and Cognition

PCA analysis estimated PC 1 to explain approximately 62 % of the variance across the nine measures of cognitive performance, PC 2 – 9 %, and PC 3 – 7 %. We therefore focus on the relationship between genetic status, RSFA, and PC 1 as a proxy for cognitive function. The cognitive variables that loaded most prominently on this component included the Trail Making Test Parts A and B, Digit Symbol Task, and Verbal Fluency, suggesting that PC 1 captures most strongly the cognitive domain of executive function. More detailed information about each PC, with explained variance and corresponding coefficients, is provided in Appendix A, Table A.6, and Figures A.2 and A.3, respectively. Kruskal-Wallis test showed a statistically significant difference in PC 1 subject scores between genetic status groups (χ2(2) = 256.02, *p* < 0.001). As anticipated, post hoc Mann-Whitney tests confirmed significantly lower PC 1 subject scores, indicative of lower cognitive function, in symptomatic carriers compared to both pre-symptomatic carriers (*U* = 1461, *p* < 0.001) and non-carriers (*U* = 1182, *p* < 0.001). No significant difference was observed between pre-symptomatic carriers and non-carriers (*U* = 36 318, *p* = 0.480).

Further regression analysis revealed a positive relationship between RSFA and cognitive function, specifically in IC 23, suggesting that individuals with higher CVR levels in left PFC performed better overall on a range of cognitive tests. In addition, a genetic status x RSFA interaction was observed in left PFC (IC 23), as well as in posterior parietal association areas (IC 17) and right lateral PFC (IC 21). The interaction effects, presented in Figure 4, highlight a stronger positive relationship between RSFA and cognitive function in mutation carriers, particularly in symptomatic individuals, than in non-carriers. ROIs analysis was overall consistent with component-based analysis (Figure 4). The output from the MLR models assessing cognitive function in relation to RSFA indices across the sample and sub-groups of interest can be consulted in Table 4.

**Figure 4.**
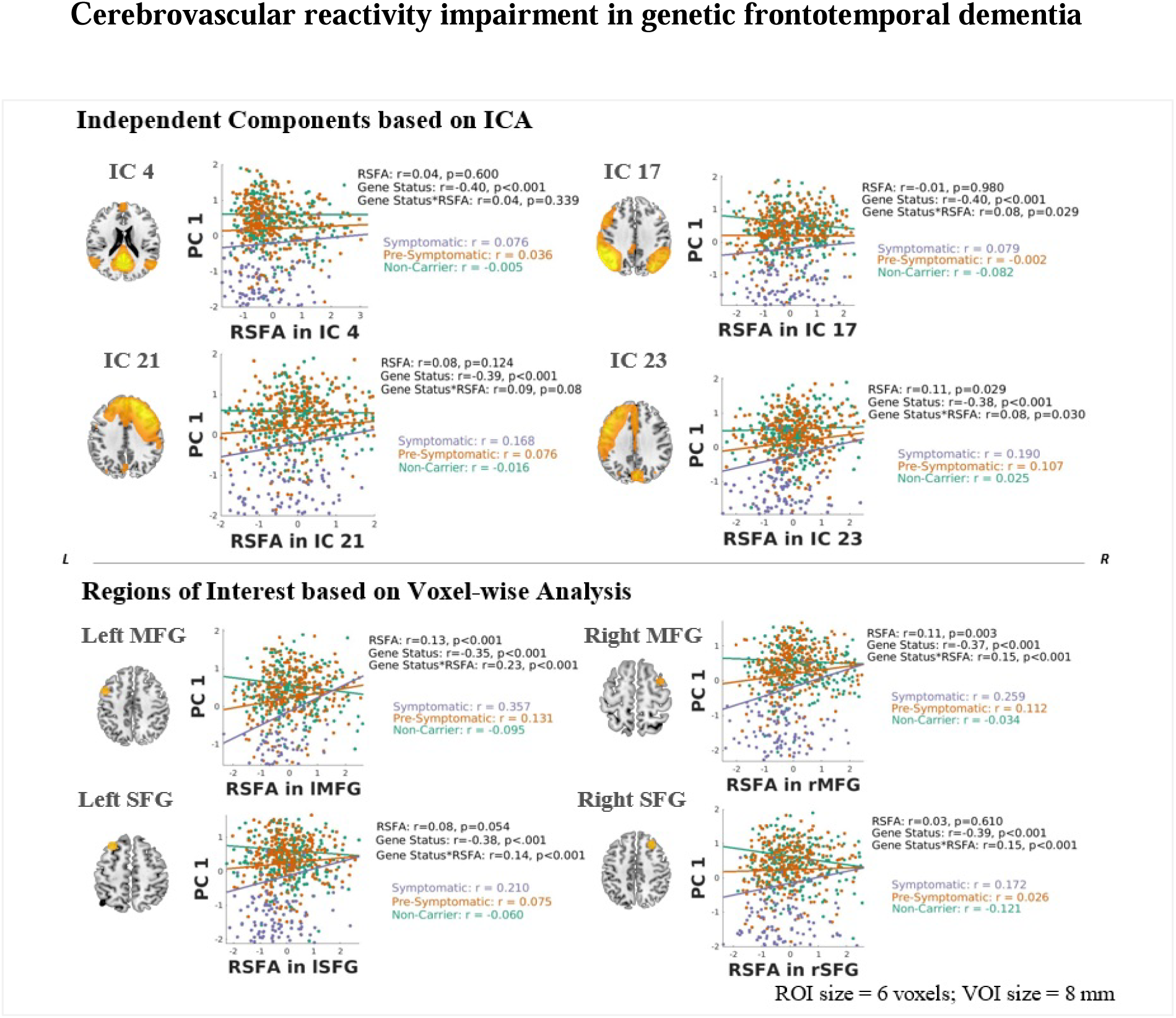
Differences in cognitive function in association with genetic status, RSFA, and genetic status x RSFA interaction across groups of interest. Cognitive function is denoted by subjects’ loading values for PC 1 following PCA on nine cognitive measures. Effects are illustrated for ICA-based components within GM areas (top panel) and several representative ROIs based on voxel-wise analysis (bottom panel). Robust general linear model regression lines for each respective IC and ROI are presented in scatter plots with corresponding r values on the right side of a representative slice depicting each IC/ROI map. P values are FDR-corrected at the 0.05 level across the whole sample. MFG, middle frontal gyrus; SFG, superior frontal gyrus. ROI, region of interest; VOI, volume of interest.

**Table 4.**
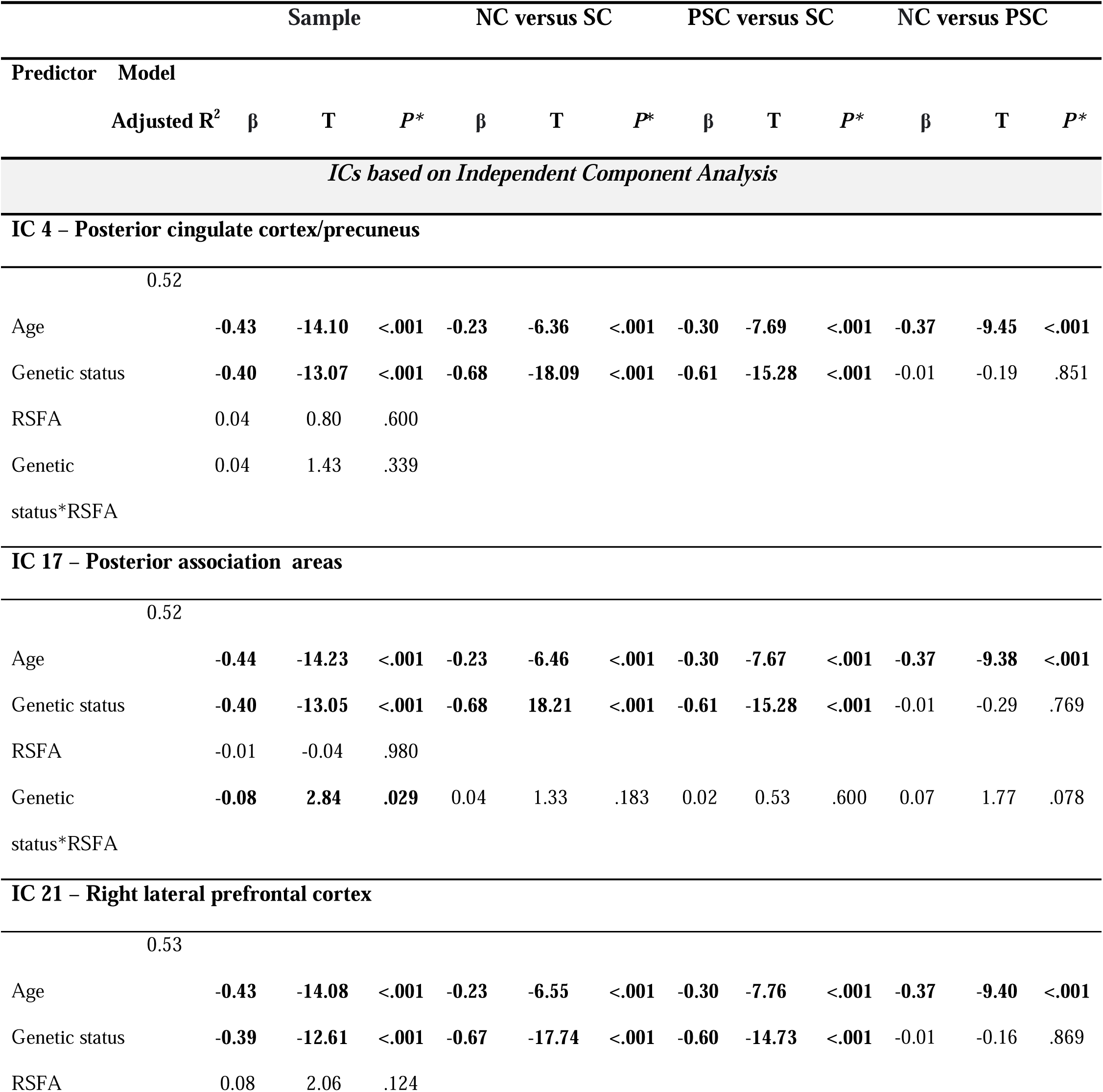
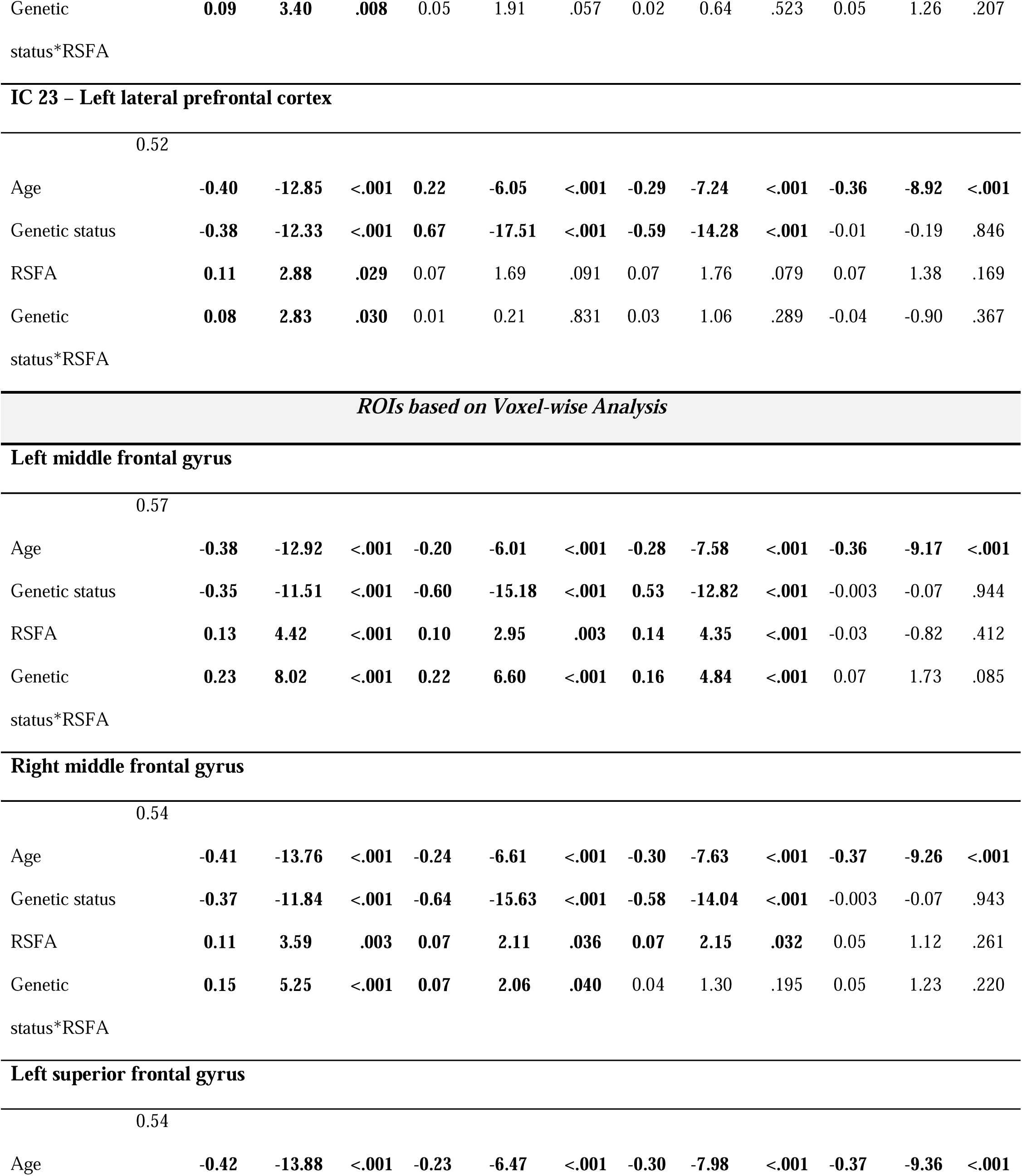
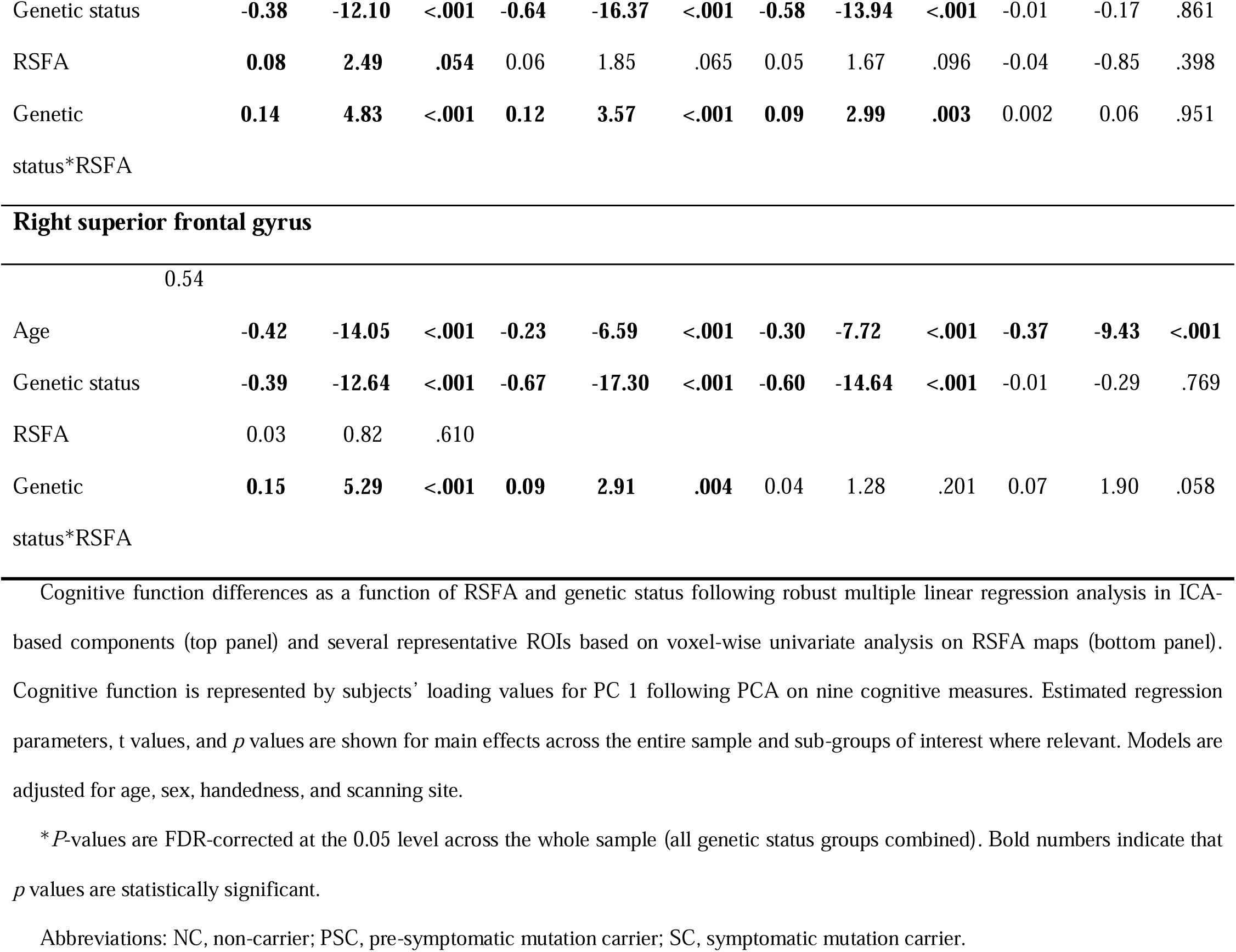
Multiple regression analysis results of cognition as a function of RSFA (model *‘CognitionPC1 ∼ 1 + Genetic status*RSFA_IC/Voxel_ + Age + Sex + Handedness + Scanning Site’)*

## 4. Discussion

We confirmed that cerebrovascular function, as measured by the resting-state fluctuation amplitudes, is reduced by mutations associated with frontotemporal dementia even in the long pre-symptomatic period. The RSFA differences worsened with disease progression and correlated with cognition in mutation carriers, over and above the effects of ageing. We propose that cerebrovascular dysfunction in genetic FTD represents an early dysregulated feature in the disease’s pathophysiology, which may interact with neurodegenerative changes.

### 4.1. Regional Distribution of Cerebrovascular Reactivity Impairment in FTD

Progressive reductions in RSFA were observed in mutation carriers versus non-carriers in ventromedial and dorsolateral prefrontal cortical areas, cingulate and parietal cortex. Comparable CVR decreases using the RSFA approach are reported in healthy ageing and acute conditions of microvascular impairment, particularly in prefrontal and superior-parietal cortical areas [28, 29, 33] that are vulnerable to lower cerebral blood flow [26, 53, 54] and the principal FTD-specific pathological burden. This regional vulnerability aligns with the observation of abnormal vasoreactivity in the default mode network (DMN) in AD [18, 19, 32, 55]. In genetic FTD, we found consistent CVR reductions in frontal cortex, anterior cingulate, and insula – regions accordant with atrophy [2, 35, 56, 57] and cerebral perfusion decreases [8, 9, 58] in pre-symptomatic and symptomatic carriers. These areas are part of the salience network, which underlies cognitive, sensory, and affective regulation, language, motor control, and social conduct, each functionally impaired in symptomatic FTD [59, 60].

We argue that the observed cerebrovascular dysfunction in FTD represents an early dysregulated pathophysiology, interacting with regional neurodegenerative changes, as postulated in AD [61–63]. Potential causes for the CVR decreases include pH dysregulation and impaired modulation of nitric oxide, which may diminish endothelium-dependent dilator responses and the dynamic range of the BOLD signal [16, 64, 65]. Alterations in the neurovascular unit, including dysfunctional vascular endothelium, hypercontractile vascular smooth muscle cells [62], depleted pericytes [5], and activated microglia [6] have been documented in familial FTD. Given the close interrelatedness between neurons and cerebral microvessels, such changes likely compromise the function of the blood-brain barrier, diminish brain perfusion, and trigger the aggregation of aberrant circulating proteins and secretion of pro-inflammatory factors, accelerating neurodegeneration [61, 62]. These findings underscore the need for further research to discern the relationship between cerebrovascular dysfunction and neurodegenerative processes in FTD and other neurodegenerative pathologies.

While the RSFA variances in inferior frontal, parietal, and precuneus (IC 4 and IC 17) accord with FTD-related hypoperfusion and atrophy profiles, the notable RSFA reductions in dorsolateral prefrontal cortex (IC 21 and IC 23) are intriguing. These regions are not commonly associated with hypoperfusion and atrophy in early FTD. This discrepancy implies a shared pathway leading to CVR impairment, hypoperfusion, and atrophy in inferior frontal and parietal regions. However, the mechanisms underlying the distinct CVR effects in dorsolateral frontal regions in FTD and the processes that interact with these changes prompt further investigation.

Such CVR alterations develop in the long pre-symptomatic window and increase with age in mutation carriers faster than in non-carriers. This suggests less effective dampening of arterial pressure pulsations through the vascular tree (i.e., diminished Windkessel effect) owing to increased arterial stiffening [26], which could influence the BOLD signal fluctuation in neighbouring tissue, including WM and CSF [66]. Such an interpretation supports previously reported RSFA increases near cerebral ventricles and vascular territories, and likely reflects the cardiovascular contribution to the RSFA signal in ageing [26, 28]. It is also plausible that the RSFA signal in ICA-identified regions captures multiple sources with different aetiology, particularly at boundaries of large vessels and adjacent perivascular space, that may exhibit different spontaneous brain activity at rest [67]. The latter illustrates the challenge of dissociating spatially overlapping sources of signal using univariate methods and motivate the use of data-driven and multimodal approaches [68], as underscored by our findings.

Although we observed diminished CVR in signature FTD frontal and parietal areas, no substantial RSFA decreases in temporal regions were found. Given the involvement of the temporal lobes, especially in *MAPT* mutations [8, 35, 56], this result may be due to type II error or the much smaller size of the *MAPT* group than the *C9orf72* and *GRN* groups. We compared the RSFA-IC loadings and RSFA-ROI estimates based on gene mutation but did not discover any significant between-group effects. This null result may be caused by small and unbalanced sub-groups per mutated gene but may also imply true commonalities in the vascular pathology downstream of the mutations’ molecular pathology. Previous neuroimaging studies have revealed gene mutation-specific brain changes in FTD [56, 69]. The CVR changes in frontal regions may reflect distinct mechanisms from the atrophy and perfusion alterations in temporal areas discovered in earlier FTD investigations. In line with this assumption, different CVR and CBF patterns have been documented in AD, with CVR deficits in prefrontal, anterior cingulate, and insular cortex proposed as direct indicators of vascular dysfunction, and CBF decreases in temporal and parietal cortices attributed to atrophy-related lower metabolic demand [55]. Our results could denote a similar mechanism whereby CVR impairment contributes to FTD disease progression both independently and conjointly with other pathophysiological processes.

### 4.2. Relationship between Cerebrovascular Impairment and Cognition

As a secondary objective, we examined the behavioural relevance of CVR alterations and found a relationship between RSFA reductions in mutation carriers and diminished cognitive function. This broadly confirmed the link between CVR decreases and impaired overall cognitive status, as previously shown in mild cognitive impairment (MCI) and AD [19], and conditions that may alter the risk of dementia [33]. Furthermore, CVR impairment predicts global cognitive performance independently of AD pathological markers, such as CSF-derived β-amyloid42 (Aβ42) and tau in healthy elderly and subjects with mixed Alzheimer’s and vascular cognitive impairment and dementia [70].

We observed an association between higher RSFA and better global cognitive function, especially in symptomatic mutation carriers, pronounced in prefrontal cortical areas – an effect that remained after adjusting for age and disease progression effects. This accords with evidence from ageing, AD, and FTD studies about the increased dependence of successful cognition on precisely regulated function within and between large-scale brain networks [71–74]. Furthermore, progressively stronger coupling between function and cognition is described in pre-symptomatic mutation carriers from the GENFI cohort as they approached their expected age of disease onset, in the absence of differences in cognitive performance relative to non-carriers [57]. Therefore, our observations support previous research and suggest that CVR may benefit cognition in FTD at-risk individuals.

CVR impairment in DMN regions did not correlate with cognition. This implies that the CVR changes in default network may not relate directly to the neuropathological processes or disease progression and may instead be influenced by other factors that modulate CVR, such as medications, as shown in ageing [75]. The nature of default network CVR changes and its implications for DMN suppression in neurodegeneration remains to be fully defined. However, findings on default network from fMRI BOLD studies should not be interpreted independently of cerebrovascular variations induced by physiological modulators [26].

### 4.3. Methodological Considerations and Future Directions

Several methodological remarks warrant consideration. Firstly, our design was cross-sectional, so any causal inferences about the associations remain to be addressed in longitudinal analyses. Second, several of the described effects only approached statistical significance, which could imply that the FDR multiple comparison correction was conservative. In the voxel-based analysis, no differences emerged between pre-symptomatic carriers and non-carriers. Despite that, the distribution of CVR effects in pre-symptomatic carriers resembled that of symptomatic cases, highlighting the vulnerability of the middle frontal and posterior cortical areas. Third, we recognise that RSFA-CVR is just one measure of cerebrovascular health. Previous examinations using the RSFA method have documented that RSFA relates to CBF effects, white matter hyperintensities (WMHs), and cardiovascular factors [26]. Thus, future investigations in the GENFI sample should clarify which vascular factors drive the RSFA changes reported here by adopting other means to quantify cerebrovascular function, such as resting arterial-spin labelling (ASL)-CBF and WMH burden on MRI. Another avenue for future efforts is to complement current analyses with estimates of functional and WM integrity, as well as CSF and blood markers in relation to cognitive decline [29] in a multi-modal manner [68, 75]. On a clinical level, using integrative approaches to uncover protective factors in prodromal stages of disease may improve prognosis and inform stratification procedures, future triallists, patients, and carers.

## 5. Concluding Remarks

Using the RSFA approach, we found CVR alterations in pre-symptomatic and symptomatic FTD with a pronounced frontal cortical predilection, concordant across component-based and voxel-level analyses. We also showed that higher CVR yields a cognitive benefit, especially in subjects at elevated FTD risk. These results demonstrate that RSFA can be used as a safe, tolerable, and clinically informative signal that can aid the quantification of cerebrovascular health in large-scale population studies among frail participants. We suggest that there is a vascular contribution that interacts with FTD pathology in driving disease expression and progression. Cerebrovascular health may be a potential target for biomarker identification and a modifiable factor, to mitigate against clinical deterioration in people at genetic risk of frontotemporal dementia.

## Data Availability

All data produced in the present study are available upon request to the authors.

## Acknowledgements

The authors would like to thank the participant volunteers and their families for their contribution to this research. We also thank Hamid Azimi for technical assistance, as well as all radiographers/technicians and research nurses from all research sites involved in this study for their invaluable support in data acquisition.

## Declaration of Interest

All authors have no conflicts of interest. Untreated to this there are several disclosures.

## Sources of Funding

K.A.T. was supported by Fellowship awards from the Guarantors of Brain (G101149) and Alzheimer’s Society, UK (grant number 602). J.B.R. has received funding from the Welcome Trust (103838; 220258) and is supported by the Cambridge University Centre for Frontotemporal Dementia, the Medical Research Council (MC_UU_00030/14; MR/T033371/1 ) and the National Institute for Health Research Cambridge Biomedical Research Centre (NIHR203312: BRC-1215-20014) and the Holt Fellowship. The views expressed are those of the authors and not necessarily those of the NIHR or the Department of Health and Social Care. J.C.V.S., L.C.J. and H.S. are supported by the Dioraphte Foundation grant 09-02-03-00, Association for Frontotemporal Dementias Research Grant 2009, Netherlands Organisation for Scientific Research grant HCMI 056-13-018, ZonMw Memorabel (Deltaplan Dementie, project number 733 051 042), ZonMw Onderzoeksprogramma Dementie (YOD-INCLUDED, project number 10510032120002), EU Joint Programme-Neurodegenerative Disease Research-GENFI-PROX, Alzheimer Nederland and the Bluefield Project. R.S-V. is supported by Alzheimer’s Research UK Clinical Research Training Fellowship (ARUK-CRF2017B-2) and has received funding from Fundació Marató de TV3, Spain (grant no. 20143810). C.G. received funding from EU Joint Programme-Neurodegenerative Disease Research-Prefrontals Vetenskapsrådet Dnr 529-2014-7504, EU Joint Programme-Neurodegenerative Disease Research-GENFI-PROX, Vetenskapsrådet 2019-0224, Vetenskapsrådet 2015-02926, Vetenskapsrådet 2018-02754, the Swedish FTD Inititative-Schörling Foundation, Alzheimer Foundation, Brain Foundation, Dementia Foundation and Region Stockholm ALF-project. D.G. received support from the EU Joint Programme-Neurodegenerative Disease Research and the Italian Ministry of Health (PreFrontALS) grant 733051042. R.V. has received funding from the Mady Browaeys Fund for Research into Frontotemporal Dementia. J.L. received funding for this work by the Deutsche Forschungsgemeinschaft German Research Foundation under Germany’s Excellence Strategy within the framework of the Munich Cluster for Systems Neurology (EXC 2145 SyNergy—ID 390857198). M.O. has received funding from Germany’s Federal Ministry of Education and Research (BMBF). E.F. has received funding from a Canadian Institute of Health Research grant #327387. M.M. has received funding from a Canadian Institute of Health Research operating grant and the Weston Brain Institute and Ontario Brain Institute. F.M. is supported by the Tau Consortium and has received funding from the Carlos III Health Institute (PI19/01637). J.D.R. is supported by the Bluefield Project and the National Institute for Health and Care Research University College London Hospitals Biomedical Research Centre and has received funding from an MRC Clinician Scientist Fellowship (MR/M008525/1) and a Miriam Marks Brain Research UK Senior Fellowship. Several authors of this publication (J.C.V.S., M.S., R.V., A.d.M., M.O., R.V., J.D.R.) are members of the European Reference Network for Rare Neurological Diseases (ERN-RND) - Project ID No 739510. This work was also supported by the EU Joint Programme-Neurodegenerative Disease Research GENFI-PROX grant [2019-02248; to J.D.R., M.O., B.B., C.G., J.C.V.S. and M.S. For the purpose of open access, the author has applied a CC BY public copyright licence to any Author Accepted Manuscript version arising from this submission.

JBR is a non-remunerated trustee of the Guarantors of Brain, Darwin College, and the PSP Association; he provides consultancy to Alzheimer Research UK, Asceneuron, Alector, Biogen, CuraSen, CumulusNeuro, UCB, SV Health, and Wave, and has research grants from AZ-Medimmune, Janssen, Lilly as industry partners in the Dementias Platform UK.

## Notes

### Competing Interest Statement

The authors have declared no competing interest.

### Funding Statement

K.A.T. was supported by Fellowship awards from the Guarantors of Brain (G101149) and Alzheimers Society, UK (grant number 602). J.B.R. has received funding from the Welcome Trust (103838; 220258) and is supported by the Cambridge University Centre for Frontotemporal Dementia, the Medical Research Council (MC_UU_00030/14; MR/T033371/1) and the National Institute for Health Research Cambridge Biomedical Research Centre (NIHR203312: BRC-1215-20014) and the Holt Fellowship. The views expressed are those of the authors and not necessarily those of the NIHR or the Department of Health and Social Care. J.C.V.S., L.C.J. and H.S. are supported by the Dioraphte Foundation grant 09-02-03-00, Association for Frontotemporal Dementias Research Grant 2009, Netherlands Organisation for Scientific Research grant HCMI 056-13-018, ZonMw Memorabel (Deltaplan Dementie, project number 733 051 042), ZonMw Onderzoeksprogramma Dementie (YOD-INCLUDED, project number 10510032120002), EU Joint Programme-Neurodegenerative Disease Research-GENFI-PROX, Alzheimer Nederland and the Bluefield Project. R.S-V. is supported by Alzheimers Research UK Clinical Research Training Fellowship (ARUK-CRF2017B-2) and has received funding from Fundacio Marato de TV3, Spain (grant no. 20143810). C.G. received funding from EU Joint Programme-Neurodegenerative Disease Research-Prefrontals Vetenskapsradet Dnr 529-2014-7504, EU Joint Programme-Neurodegenerative Disease Research-GENFI-PROX, Vetenskapsradet 2019-0224, Vetenskapsradet 2015-02926, Vetenskapsradet 2018-02754, the Swedish FTD Inititative-Schorling Foundation, Alzheimer Foundation, Brain Foundation, Dementia Foundation and Region Stockholm ALF-project. D.G. received support from the EU Joint Programme Neurodegenerative Disease Research and the Italian Ministry of Health (PreFrontALS) grant 733051042. R.V. has received funding from the Mady Browaeys Fund for Research into Frontotemporal Dementia. J.L. received funding for this work by the Deutsche Forschungsgemeinschaft German Research Foundation under Germany Excellence Strategy within the framework of the Munich Cluster for Systems Neurology (EXC 2145 SyNergy ID 390857198). M.O. has received funding from Ministry of Education and Research (BMBF) in Germany. E.F. has received funding from a Canadian Institute of Health Research grant number 327387. M.M. has received funding from a Canadian Institute of Health Research operating grant and the Weston Brain Institute and Ontario Brain Institute. F.M. is supported by the Tau Consortium and has received funding from the Carlos III Health Institute (PI19/01637). J.D.R. is supported by the Bluefield Project and the National Institute for Health and Care Research University College London Hospitals Biomedical Research Centre and has received funding from an MRC Clinician Scientist Fellowship (MR/M008525/1) and a Miriam Marks Brain Research UK Senior Fellowship. Several authors of this publication (J.C.V.S., M.S., R.V., A.d.M., M.O., R.V., J.D.R.) are members of the European Reference Network for Rare Neurological Diseases (ERN-RND) Project ID No 739510. This work was also supported by the EU Joint Programme-Neurodegenerative Disease Research GENFI-PROX grant [2019-02248; to J.D.R., M.O., B.B., C.G., J.C.V.S. and M.S. For the purpose of open access, the author has applied a CC BY public copyright licence to any Author Accepted Manuscript version arising from this submission.
JBR is a non-remunerated trustee of the Guarantors of Brain, Darwin College, and the PSP Association; he provides consultancy to Alzheimer Research UK, Asceneuron, Alector, Biogen, CuraSen, CumulusNeuro, UCB, SV Health, and Wave, and has research grants from AZ-Medimmune, Janssen, Lilly as industry partners in the Dementias Platform UK.

### Author Declarations

The study uses data from human patients from the Genetic Frontotemporal Dementia (GENFI) Consortium initiative. The data are available upon request from the respective consortium website. (https://www.genfi.org/)

## References

1. Coyle-Gilchrist IT, Dick KM, Patterson K, Vázquez Rodríquez P, Wehmann E, Wilcox A, Lansdall CJ, Dawson KE, Wiggins J, Mead S, Brayne C. Prevalence, characteristics, and survival of frontotemporal lobar degeneration syndromes. Neurology. 2016 May 3;86(18):1736–43. 10.1007/S00415-019-09363-4

2. Greaves CV, Rohrer JD. An update on genetic frontotemporal dementia. Journal of neurology. 2019 Aug 1;266(8):2075–86. 10.1007/s00415-019-09363-4

3. Meeter LH, Kaat LD, Rohrer JD, Van Swieten JC. Imaging and fluid biomarkers in frontotemporal dementia. Nature Reviews Neurology. 2017 Jul;13(7):406–19. 10.1038/nrneurol.2017.75

4. Raz L, Knoefel J, Bhaskar K. The neuropathology and cerebrovascular mechanisms of dementia. Journal of Cerebral Blood Flow & Metabolism. 2016 Jan;36(1):172–86. 10.1038/jcbfm.2015.164

5. Gerrits E, Giannini LA, Brouwer N, Melhem S, Seilhean D, Le Ber I, Brainbank Neuro-CEB Neuropathology Network, Kamermans A, Kooij G, de Vries HE, Boddeke EW. Neurovascular dysfunction in GRN-associated frontotemporal dementia identified by single-nucleus RNA sequencing of human cerebral cortex. Nature neuroscience. 2022 Aug;25(8):1034–48. 10.1038/s41593-022-01124-3

6. Malpetti M, Rittman T, Jones PS, Cope TE, Passamonti L, Bevan-Jones WR, Patterson K, Fryer TD, Hong YT, Aigbirhio FI, O’Brien JT. In vivo PET imaging of neuroinflammation in familial frontotemporal dementia. Journal of Neurology, Neurosurgery & Psychiatry. 2020 Oct 29. 10.1136/jnnp-2020-323698

7. Du AT, Jahng GH, Hayasaka S, Kramer JH, Rosen HJ, Gorno-Tempini ML, Rankin KP, Miller BL, Weiner MW, Schuff N. Hypoperfusion in frontotemporal dementia and Alzheimer disease by arterial spin labeling MRI. Neurology. 2006 Oct 10;67(7):1215–20.

8. Mutsaerts HJ, Mirza SS, Petr J, Thomas DL, Cash DM, Bocchetta M, De Vita E, Metcalfe AW, Shirzadi Z, Robertson AD, Tartaglia MC. Cerebral perfusion changes in presymptomatic genetic frontotemporal dementia: a GENFI study. Brain. 2019 Apr 1;142(4):1108–20. 10.1093/brain/awz039

9. Dopper EG, Chalos V, Ghariq E, den Heijer T, Hafkemeijer A, Jiskoot LC, de Koning I, Seelaar H, van Minkelen R, van Osch MJ, Rombouts SA. Cerebral blood flow in presymptomatic MAPT and GRN mutation carriers: a longitudinal arterial spin labeling study. NeuroImage: Clinical. 2016 Feb 1;12:460–5. 10.1016/j.nicl.2016.08.001

10. Thal DR, von Arnim CA, Griffin WS, Mrak RE, Walker L, Attems J, Arzberger T. Frontotemporal lobar degeneration FTLD-tau: preclinical lesions, vascular, and Alzheimer-related co-pathologies. Journal of neural transmission. 2015 Jul;122:1007–18. 10.1007/s00702-014-1360-6

11. Willie CK, Tzeng YC, Fisher JA, Ainslie PN. Integrative regulation of human brain blood flow. The Journal of physiology. 2014 Mar 1;592(5):841–59. 10.1113/jphysiol.2013.268953

12. Jensen KE, Thomsen C, Henriksen O. In vivo measurement of intracellular pH in human brain during different tensions of carbon dioxide in arterial blood. A31PLJNMR study. Acta physiologica scandinavica. 1988 Oct;134(2):295-8. 10.1111/j.1748-1716.1988.tb08492.x

13. Lassen NA. Brain extracellular pH: the main factor controlling cerebral blood flow. Scandinavian journal of clinical and laboratory investigation. 1968 Jan 1;22(4):247–51.

14. Reich T, Rusinek H. Cerebral cortical and white matter reactivity to carbon dioxide. Stroke. 1989 Apr;20(4):453–7. 10.1161/01.STR.20.4.453

15. Tsuda Y, Hartmann A. Changes in hyperfrontality of cerebral blood flow and carbon dioxide reactivity with age. Stroke. 1989 Dec;20(12):1667–73. 10.1161/01.STR.20.12.1667

16. Brandes RP, Fleming I, Busse R. Endothelial aging. Cardiovascular research. 2005 May 1;66(2):286-94. 10.1016/j.cardiores.2004.12.027

17. Haight TJ, Bryan RN, Erus G, Davatzikos C, Jacobs DR, D’Esposito M, Lewis CE, Launer LJ. Vascular risk factors, cerebrovascular reactivity, and the default-mode brain network. Neuroimage. 2015 Jul 15;115:7–16. 10.1016/j.neuroimage.2015.04.039

18. Cantin S, Villien M, Moreaud O, Tropres I, Keignart S, Chipon E, Le Bas JF, Warnking J, Krainik A. Impaired cerebral vasoreactivity to CO2 in Alzheimer’s disease using BOLD fMRI. Neuroimage. 2011 Sep 15;58(2):579–87. 10.1016/j.neuroimage.2011.06.070

19. Richiardi J, Monsch AU, Haas T, Barkhof F, Van de Ville D, Radü EW, Kressig RW, Haller S. Altered cerebrovascular reactivity velocity in mild cognitive impairment and Alzheimer’s disease. Neurobiology of aging. 2015 Jan 1;36(1):33–41. 10.1016/j.neurobiolaging.2014.07.020

20. Birn RM, Diamond JB, Smith MA, Bandettini PA. Separating respiratory-variation-related fluctuations from neuronal-activity-related fluctuations in fMRI. Neuroimage. 2006 Jul 15;31(4):1536–48. 10.1016/j.neuroimage.2006.02.048

21. Shmueli K, van Gelderen P, de Zwart JA, Horovitz SG, Fukunaga M, Jansma JM, Duyn JH. Low-frequency fluctuations in the cardiac rate as a source of variance in the resting-state fMRI BOLD signal. Neuroimage. 2007 Nov 1;38(2):306–20.

22. Shmueli K, van Gelderen P, de Zwart JA, Horovitz SG, Fukunaga M, Jansma JM, Duyn JH. Low-frequency fluctuations in the cardiac rate as a source of variance in the resting-state fMRI BOLD signal. Neuroimage. 2007 Nov 1;38(2):306–20. 10.1016/j.neuroimage.2007.07.037

22. Golestani AM, Wei LL, Chen JJ. Quantitative mapping of cerebrovascular reactivity using resting-state BOLD fMRI: validation in healthy adults. Neuroimage. 2016 Sep 1;138:147–63. 10.1016/j.neuroimage.2016.05.025

23. Kannurpatti SS, Biswal BB. Detection and scaling of task-induced fMRI-BOLD response using resting state fluctuations. Neuroimage. 2008 May 1;40(4):1567–74. 10.1016/j.neuroimage.2007.09.040

24. Liu P, Li Y, Pinho M, Park DC, Welch BG, Lu H. Cerebrovascular reactivity mapping without gas challenges. Neuroimage. 2017 Feb 1;146:320–6.

25. Rostrup E, Larsson HB, Toft PB, Garde K, Thomsen C, Ring P, Søndergaard L, Henriksen O. Functional MRI of CO2 induced increase in cerebral perfusion. NMR in Biomedicine. 1994 Mar;7(1LJ2):29-34. 10.1002/nbm.1940070106

26. Tsvetanov KA, Henson RN, Rowe JB. Separating vascular and neuronal effects of age on fMRI BOLD signals. Philosophical Transactions of the Royal Society B. 2021 Jan 4;376(1815):20190631. 10.1098/rstb.2019.0631

27. Kannurpatti SS, Motes MA, Biswal BB, Rypma B. Assessment of unconstrained cerebrovascular reactivity marker for large age-range FMRI studies. PloS one. 2014 Feb 13;9(2):e88751. 10.1371/journal.pone.0088751

28. Tsvetanov KA, Henson RN, Tyler LK, Davis SW, Shafto MA, Taylor JR, Williams N, Rowe JB. The effect of ageing on fMRI: Correction for the confounding effects of vascular reactivity evaluated by joint fMRI and MEG in 335 adults. Human brain mapping. 2015 Jun;36(6):2248–69. 10.1002/hbm.22768

29. Tsvetanov KA, Henson RN, Jones PS, Mutsaerts H, Fuhrmann D, Tyler LK, CamLJCAN, Rowe JB. The effects of age on restingLJstate BOLD signal variability is explained by cardiovascular and cerebrovascular factors. Psychophysiology. 2021 Jul;58(7):e13714. 10.1111/psyp.13714

30. Su J, Wang M, Ban S, Wang L, Cheng X, Hua F, Tang Y, Zhou H, Zhai Y, Du X, Liu J. Relationship between changes in resting-state spontaneous brain activity and cognitive impairment in patients with CADASIL. The Journal of Headache and Pain. 2019 Dec;20:1–1. 10.1186/s10194-019-0982-3

31. Nair VA, Raut RV, Prabhakaran V. Investigating the blood oxygenation level-dependent functional MRI response to a verbal fluency task in early stroke before and after hemodynamic scaling. Frontiers in Neurology. 2017 Jun 19;8:283. 10.3389/fneur.2017.00283

32. Millar PR, Ances BM, Gordon BA, Benzinger TL, Fagan AM, Morris JC, Balota DA. Evaluating resting-state BOLD variability in relation to biomarkers of preclinical Alzheimer’s disease. Neurobiology of aging. 2020 Dec 1;96:233–45. 10.1016/j.neurobiolaging.2020.08.007

33. Tsvetanov KA, Spindler LR, Stamatakis EA, Newcombe VF, Lupson VC, Chatfield DA, Manktelow AE, Outtrim JG, Elmer A, Kingston N, Bradley JR. Hospitalisation for COVID-19 predicts long lasting cerebrovascular impairment: A prospective observational cohort study. NeuroImage: Clinical. 2022 Jan 1;36:103253. 10.1016/j.nicl.2022.103253

34. Morris JC, Weintraub S, Chui HC, Cummings J, DeCarli C, Ferris S, Foster NL, Galasko D, Graff-Radford N, Peskind ER, Beekly D. The Uniform Data Set (UDS): clinical and cognitive variables and descriptive data from Alzheimer Disease Centers. Alzheimer Disease & Associated Disorders. 2006 Oct 1;20(4):210–6. 10.1097/01.wad.0000213865.09806.92

35. Rohrer JD, Nicholas JM, Cash DM, Van Swieten J, Dopper E, Jiskoot L, Van Minkelen R, Rombouts SA, Cardoso MJ, Clegg S, Espak M. Presymptomatic cognitive and neuroanatomical changes in genetic frontotemporal dementia in the Genetic Frontotemporal dementia Initiative (GENFI) study: a cross-sectional analysis. The Lancet Neurology. 2015 Mar 1;14(3):253–62. 10.1016/S1474-4422(14)70324-2

36. Van Buuren S, Groothuis-Oudshoorn K. mice: Multivariate imputation by chained equations in R. Journal of statistical software. 2011 Dec 12;45:1–67. 10.18637/jss.v045.i03

37. Jenkinson M, Beckmann CF, Behrens TE, Woolrich MW, Smith SM. Fsl. Neuroimage. 2012 Aug 15;62(2):782-90. 10.1016/j.neuroimage.2011.09.015

38. Smith SM, Jenkinson M, Woolrich MW, Beckmann CF, Behrens TE, Johansen-Berg H, Bannister PR, De Luca M, Drobnjak I, Flitney DE, Niazy RK. Advances in functional and structural MR image analysis and implementation as FSL. Neuroimage. 2004 Jan 1;23:S208–19. 10.1016/j.neuroimage.2004.07.051

39. Penny WD, Friston KJ, Ashburner JT, Kiebel SJ, Nichols TE, editors. Statistical parametric mapping: the analysis of functional brain images. Elsevier; 2011 Apr 28.

40. Jenkinson M, Bannister P, Brady M, Smith S. Improved optimization for the robust and accurate linear registration and motion correction of brain images. Neuroimage. 2002 Oct 1;17(2):825–41. 10.1006/nimg.2002.1132

41. Pruim RH, Mennes M, van Rooij D, Llera A, Buitelaar JK, Beckmann CF. ICA-AROMA: A robust ICA-based strategy for removing motion artifacts from fMRI data. Neuroimage. 2015 May 15;112:267–77. 10.1016/j.neuroimage.2015.02.064

42. Geerligs L, Tsvetanov KA, Henson RN. Challenges in measuring individual differences in functional connectivity using fMRI: the case of healthy aging. Human brain mapping. 2017 Aug;38(8):4125–56. 10.1002/hbm.23653

43. Satterthwaite TD, Elliott MA, Gerraty RT, Ruparel K, Loughead J, Calkins ME, Eickhoff SB, Hakonarson H, Gur RC, Gur RE, Wolf DH. An improved framework for confound regression and filtering for control of motion artifact in the preprocessing of resting-state functional connectivity data. Neuroimage. 2013 Jan 1;64:240–56. 10.1016/j.neuroimage.2012.08.052

44. Xu L, Groth KM, Pearlson G, Schretlen DJ, Calhoun VD. SourceLJbased morphometry: The use of independent component analysis to identify gray matter differences with application to schizophrenia. Human brain mapping. 2009 Mar;30(3):711–24. 10.1002/hbm.20540

45. Hui M, Li J, Wen X, Yao L, Long Z. An empirical comparison of information-theoretic criteria in estimating the number of independent components of fMRI data. PloS one. 2011 Dec 27;6(12):e29274. 10.1371/journal.pone.0029274

46. Li YO, Adalı T, Calhoun VD. Estimating the number of independent components for functional magnetic resonance imaging data. Human brain mapping. 2007 Nov;28(11):1251–66. 10.1002/hbm.20359

47. Rissanen J. Modeling by shortest data description. Automatica. 1978 Sep 1;14(5):465–71. 10.1016/0005-1098(78)90005-5

48. Himberg J, Hyvärinen A, Esposito F. Validating the independent components of neuroimaging time series via clustering and visualization. Neuroimage. 2004 Jul 1;22(3):1214–22. 10.1016/j.neuroimage.2004.03.027

49. Millar PR, Petersen SE, Ances BM, Gordon BA, Benzinger TL, Morris JC, Balota DA. Evaluating the sensitivity of resting-state BOLD variability to age and cognition after controlling for motion and cardiovascular influences: a network-based approach. Cerebral Cortex. 2020 Nov;30(11):5686–701. 10.1093/cercor/bhaa138

50. Moore KM, Nicholas J, Grossman M, McMillan CT, Irwin DJ, Massimo L, Van Deerlin VM, Warren JD, Fox NC, Rossor MN, Mead S. Age at symptom onset and death and disease duration in genetic frontotemporal dementia: an international retrospective cohort study. The Lancet Neurology. 2020 Feb 1;19(2):145–56. 10.1016/S1474-4422(19)30394-1

51. Chen J, Liu J, Calhoun VD, Arias-Vasquez A, Zwiers MP, Gupta CN, Franke B, Turner JA. Exploration of scanning effects in multi-site structural MRI studies. Journal of neuroscience methods. 2014 Jun 15;230:37–50. 10.1016/j.jneumeth.2014.04.023

52. Faria AV, Joel SE, Zhang Y, Oishi K, van Zjil PC, Miller MI, Pekar JJ, Mori S. Atlas-based analysis of resting-state functional connectivity: Evaluation for reproducibility and multi-modal anatomy–function correlation studies. Neuroimage. 2012 Jul 2;61(3):613–21. 10.1016/j.neuroimage.2012.03.078

53. Flück D, Beaudin AE, Steinback CD, Kumarpillai G, Shobha N, McCreary CR, Peca S, Smith EE, Poulin MJ. Effects of aging on the association between cerebrovascular responses to visual stimulation, hypercapnia and arterial stiffness. Frontiers in physiology. 2014 Feb 19;5:49. 10.3389/fphys.2014.00049

54. Hosp JA, Dressing A, Blazhenets G, Bormann T, Rau A, Schwabenland M, Thurow J, Wagner D, Waller C, Niesen WD, Frings L. Cognitive impairment and altered cerebral glucose metabolism in the subacute stage of COVID-19. Brain. 2021 Apr 1;144(4):1263–76.

55. Yezhuvath US, Uh J, Cheng Y, Martin-Cook K, Weiner M, Diaz-Arrastia R, van Osch M, Lu H. Forebrain-dominant deficit in cerebrovascular reactivity in Alzheimer’s disease. Neurobiology of aging. 2012 Jan 1;33(1):75–82. 10.1016/j.neurobiolaging.2010.02.005

56. Cash DM, Bocchetta M, Thomas DL, Dick KM, van Swieten JC, Borroni B, Galimberti D, Masellis M, Tartaglia MC, Rowe JB, Graff C. Patterns of gray matter atrophy in genetic frontotemporal dementia: results from the GENFI study. Neurobiology of aging. 2018 Feb 1;62:191–6. 10.1016/j.neurobiolaging.2017.10.008

57. Tsvetanov KA, Gazzina S, Jones PS, van Swieten J, Borroni B, SanchezLJValle R, Moreno F, Laforce Jr R, Graff C, Synofzik M, Galimberti D. Brain functional network integrity sustains cognitive function despite atrophy in presymptomatic genetic frontotemporal dementia. Alzheimer’s & Dementia. 2021 Mar;17(3):500–14. 10.1002/alz.12209

58. Mutsaerts HJ, Petr J, Thomas DL, De Vita E, Cash DM, van Osch MJ, Golay X, Groot PF, Ourselin S, van Swieten J, Laforce Jr R. Comparison of arterial spin labeling registration strategies in the multiLJcenter GENetic frontotemporal dementia initiative (GENFI). Journal of Magnetic Resonance Imaging. 2018 Jan;47(1):131–40. 10.1002/jmri.25751

59. Menon V, Uddin LQ. Saliency, switching, attention and control: a network model of insula function. Brain structure and function. 2010 Jun;214:655–67. 10.1007/s00429-010-0262-0

60. Seeley WW. Anterior insula degeneration in frontotemporal dementia. Brain Structure and Function. 2010 Jun;214:465–75. 10.1007/s00429-010-0263-z

61. De la Torre JC. Alzheimer disease as a vascular disorder: nosological evidence. Stroke. 2002 Apr 1;33(4):1152–62. 10.1161/01.STR.0000014421.15948.67

62. Iadecola C. Neurovascular regulation in the normal brain and in Alzheimer’s disease. Nature Reviews Neuroscience. 2004 May 1;5(5):347–60. 10.1038/nrn1387

63. Chun MY, Jang H, Kim SJ, Park YH, Yun J, Lockhart SN, Weiner M, De Carli C, Moon SH, Choi JY, Nam KR. Emerging role of vascular burden in AT (N) classification in individuals with Alzheimer’s and concomitant cerebrovascular burdens. Journal of Neurology, Neurosurgery & Psychiatry. 2024 Jan 1;95(1):44–51. 10.1136/jnnp-2023-331603

64. Claassen JA, Jansen RW. Cholinergically mediated augmentation of cerebral perfusion in alzheimer’s disease and related cognitive disorders: the cholinergic– vascular hypothesis. The Journals of Gerontology Series A: Biological Sciences and Medical Sciences. 2006 Mar 1;61(3):267–71. 10.1093/gerona/61.3.267

65. Lyons D, Roy S, Patel M, Benjamin N, Swift CG. Impaired nitric oxide-mediated vasodilatation and total body nitric oxide production in healthy old age. Clinical Science. 1997 Dec 1;93(6):519–25. 10.1042/cs0930519

66. Makedonov I, Black SE, MacIntosh BJ. BOLD fMRI in the white matter as a marker of aging and small vessel disease. PloS one. 2013 Jul 2;8(7):e67652. 10.1371/journal.pone.0067652

67. Premi E, Calhoun VD, Diano M, Gazzina S, Cosseddu M, Alberici A, Archetti S, Paternicò D, Gasparotti R, van Swieten J, Galimberti D. The inner fluctuations of the brain in presymptomatic frontotemporal dementia: the chronnectome fingerprint. Neuroimage. 2019 Apr 1;189:645–54. 10.1016/j.neuroimage.2019.01.080

68. Liu X, Tyler LK, CamLJCAN, Rowe JB, Tsvetanov KA. Multimodal fusion analysis of functional, cerebrovascular, and structural neuroimaging in healthy aging subjects. Human brain mapping. 2022 Dec 15;43(18):5490–508. 10.1002/hbm.26025

69. Rohrer JD, Ridgway GR, Modat M, Ourselin S, Mead S, Fox NC, Rossor MN, Warren JD. Distinct profiles of brain atrophy in frontotemporal lobar degeneration caused by progranulin and tau mutations. Neuroimage. 2010 Nov 15;53(3):1070–6. 10.1016/j.neuroimage.2009.12.088

70. Sur S, Lin Z, Li Y, Yasar S, Rosenberg P, Moghekar A, Hou X, Kalyani R, Hazel K, Pottanat G, Xu C. Association of cerebrovascular reactivity and Alzheimer pathologic markers with cognitive performance. Neurology. 2020 Aug 25;95(8):e962–72. 10.1212/WNL.0000000000010133

71. Liu X, Tyler LK, Davis SW, Rowe JB, Tsvetanov KA. Cognition’s dependence on functional network integrity with age is conditional on structural network integrity. Neurobiology of Aging. 2023 Sep 1;129:195–208. 10.1016/j.neurobiolaging.2023.06.001

72. Passamonti L, Tsvetanov KA, Jones PS, Bevan-Jones WR, Arnold R, Borchert RJ, Mak E, Su L, O’brien JT, Rowe JB. Neuroinflammation and functional connectivity in Alzheimer’s disease: interactive influences on cognitive performance. Journal of Neuroscience. 2019 Sep 4;39(36):7218–26. 10.1523/JNEUROSCI.2574-18.2019

73. Tsvetanov KA, Henson RN, Tyler LK, Razi A, Geerligs L, Ham TE, Rowe JB. Extrinsic and intrinsic brain network connectivity maintains cognition across the lifespan despite accelerated decay of regional brain activation. Journal of Neuroscience. 2016 Mar 16;36(11):3115–26. 10.1523/JNEUROSCI.2733-15.2016

74. Tsvetanov KA, Ye Z, Hughes L, Samu D, Treder MS, Wolpe N, Tyler LK, Rowe JB. Activity and connectivity differences underlying inhibitory control across the adult life span. Journal of Neuroscience. 2018 Sep 5;38(36):7887–900. 10.1523/JNEUROSCI.2919-17.2018

75. Wu S, Tyler LK, Henson RN, Rowe JB, Tsvetanov KA. Cerebral blood flow predicts multiple demand network activity and fluid intelligence across the adult lifespan. Neurobiology of aging. 2023 Jan 1;121:1–4. 10.1016/j.neurobiolaging.2022.09.006

