## Supplementary material for "Cerebrovascular reactivity impairment in genetic frontotemporal dementia": Appendix A_Kancheva et al._MedRxiv.docx

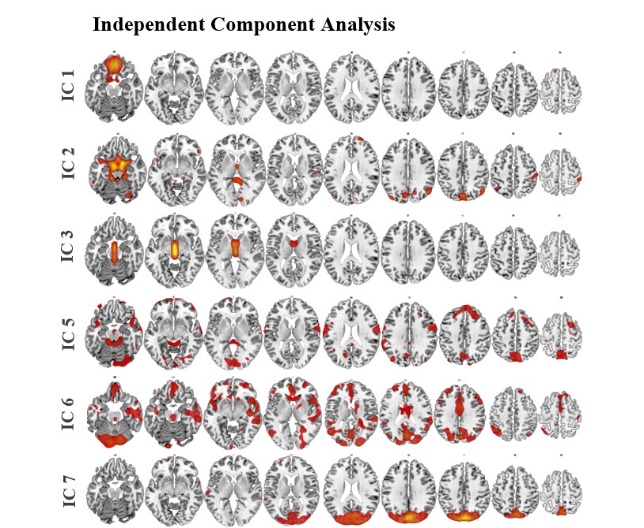
**Appendix A:**

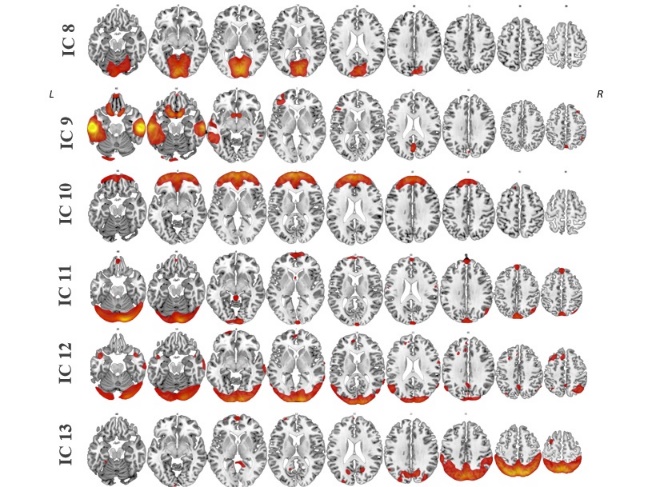

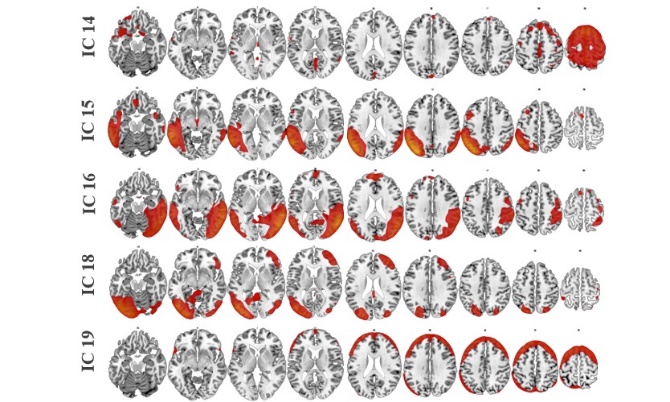

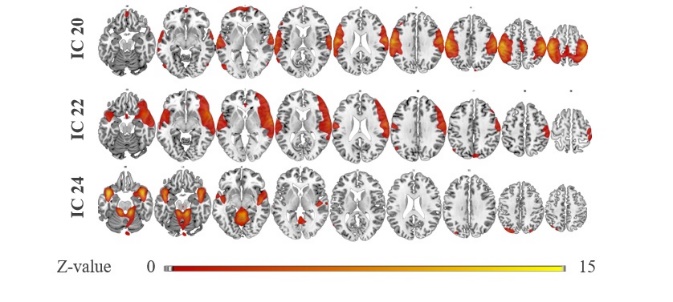

***Figure A.1.*** *Spatial distribution of 20 independent components (ICs) based on group ICA on RSFA maps across subjects. The number of components is generated according to the MDL criterion. Components show spatial distribution within vascular territories, areas in the proximity of fluid-filled ventricles, and GM regions. Group-level spatial maps are overlaid onto the Colin-27 (ch2.nii) structural template of the MNI brain, where intensity values correspond to z-values.*

Z-score	5
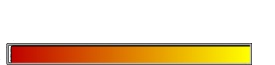
 15

| **Table A.1.** Multiple regression analysis results of independent component (IC) subject loadings from independent component analysis (model*‘RSFA_IC_ ~ 1 + Genetic Status*Age + Sex + Handedness + Scanning Site’)* | | | | | | | | | | | | | | |
| --- | --- | --- | --- | --- | --- | --- | --- | --- | --- | --- | --- | --- | --- | --- |
| **IC Model**  **adjusted R^2^ Age Genetic Status Genetic Status*Age** | | | | | | | | | | | | | | |
|  |  | **β** | **T** | ***P*** | **FDR- corr. *P*** | **β** | **T** | ***P*** | **FDR-**  **corr. *P*** | **β** | **T** | ***P*** | **FDR-**  **corr. *P*** | |
| **IC 1** |  |  |  |  |  |  |  |  |  |  |  |  | |  |
| Sample | 0.23 | 0.06 | 2.10 | 0.04 | 0.15 | 0.02 | 0.55 | 0.58 | 0.88 | -0.01 | -0.40 | 0.69 | | 0.88 |
| **IC 2** |  |  |  |  |  |  |  |  |  |  |  |  | |  |
| Sample | 0.71 | 0.04 | 1.93 | 0.05 | 0.11 | 0.003 | 0.14 | 0.89 | 0.89 | 0.05 | 2.31 | 0.02 | | 0.06 |
| **IC 3** |  |  |  |  |  |  |  |  |  |  |  |  | |  |
| Sample | 0.62 | 0.11 | 4.74 | <.001 | <.001 | 0.04 | 1.33 | 0.19 | 0.32 | 0.01 | 0.51 | 0.61 | | 0.71 |
| **IC 5** |  |  |  |  |  |  |  |  |  |  |  |  | |  |
| Sample | 0.71 | -0.05 | -1.84 | 0.07 | 0.22 | -0.01 | -0.23 | 0.82 | 0.86 | 0.01 | 0.21 | 0.83 | | 0.87 |
| **IC 6** |  |  |  |  |  |  |  |  |  |  |  |  | |  |
| Sample | 0.23 | 0.02 | 0.51 | 0.61 | 0.70 | 0.02 | 0.59 | 0.56 | 0.67 | -0.01 | -0.33 | 0.74 | | 0.81 |
| **IC 7** |  |  |  |  |  |  |  |  |  |  |  |  | |  |
| Sample | 0.37 | -0.16 | -4.68 | <.001 | <.001 | 0.05 | 1.31 | 0.19 | 0.32 | 0.02 | 0.56 | 0.58 | | 0.65 |
| **IC 8** |  |  |  |  |  |  |  |  |  |  |  |  | |  |
| Sample | 0.36 | -0.19 | -5.86 | <.001 | <.001 | -0.01 | -0.34 | 0.73 | 0.78 | 0.06 | 1.91 | 0.06 | | 0.11 |
| **IC 9** |  |  |  |  |  |  |  |  |  |  |  |  | |  |
| Sample | 0.60 | 0.06 | 2.44 | 0.01 | 0.02 | -0.01 | -0.37 | 0.71 | 0.79 | 0.002 | 0.07 | 0.94 | | 0.94 |
| **IC 10** |  |  |  |  |  |  |  |  |  |  |  |  | |  |
| Sample | 0.72 | -0.03 | -1.28 | 0.20 | 0.25 | -0.01 | -0.55 | 0.58 | 0.63 | -0.003 | -0.13 | 0.90 | | 0.93 |
| **IC 11** |  |  |  |  |  |  |  |  |  |  |  |  | |  |
| Sample | 0.17 | -0.24 | -6.42 | <.001 | <.001 | -0.03 | -0.82 | 0.41 | 0.65 | 0.07 | 1.80 | 0.07 | | 0.23 |
| **IC 12** |  |  |  |  |  |  |  |  |  |  |  |  | |  |
| Sample | 0.49 | -0.09 | -2.85 | 0.05 | 0.02 | 0.02 | 0.74 | 0.46 | 0.57 | 0.06 | 2.07 | 0.04 | | 0.11 |
| **IC 13** |  |  |  |  |  |  |  |  |  |  |  |  | |  |
| Sample | 0.36 | -0.18 | -5.04 | <.001 | <.001 | -0.02 | -0.50 | 0.62 | 0.68 | 0.001 | 0.02 | 0.99 | | 0.99 |
| **IC 14** |  |  |  |  |  |  |  |  |  |  |  |  | |  |
| Sample | 0.60 | -0.14 | -5.28 | <.001 | <.001 | -0.01 | -0.24 | 0.81 | 0.84 | -0.02 | -0.77 | 0.44 | | 0.55 |
| **IC 15** |  |  |  |  |  |  |  |  |  |  |  |  | |  |
| Sample | 0.69 | -0.04 | -1.60 | .109 | .156 | 0.06 | 2.09 | .037 | .066 | -0.03 | -1.10 | .273 | | .342 |
| **IC 16** |  |  |  |  |  |  |  |  |  |  |  |  | |  |
| Sample | 0.38 | -0.08 | -2.40 | 0.02 | 0.04 | -0.04 | -1.02 | 0.31 | 0.40 | 0.04 | 1.08 | 0.28 | | 0.40 |
| **IC 18** |  |  |  |  |  |  |  |  |  |  |  |  | |  |
| Sample | 0.26 | -0.17 | -4.44 | <.001 | <.001 | 0.06 | 1.42 | 0.16 | 0.22 | -0.002 | -0.05 | 0.96 | | 0.96 |
| **IC 19** |  |  |  |  |  |  |  |  |  |  |  |  | |  |
| Sample | 0.24 | -0.20 | -5.57 | <.001 | <.001 | -0.17 | -4.33 | <.001 | <.001 | -0.08 | -2.26 | 0.02 | | 0.09 |
| **IC 20** |  |  |  |  |  |  |  |  |  |  |  |  | |  |
| Sample | 0.54 | -0.18 | -6.29 | <.001 | <.001 | -0.01 | -0.26 | 0.80 | 0.80 | -0.02 | -0.61 | 0.54 | | 0.58 |
| **IC 22** |  |  |  |  |  |  |  |  |  |  |  |  | |  |
| Sample | 0.56 | 0.10 | 3.52 | <.001 | 0.001 | -0.02 | -0.67 | 0.50 | 0.56 | -0.02 | -0.64 | 0.52 | | 0.56 |
| **IC 24** |  |  |  |  |  |  |  |  |  |  |  |  | |  |
| Sample | 0.79 | 0.12 | 5.99 | <.001 | <.001 | 0.02 | 1.00 | 0.32 | 0.41 | 0.02 | 1.24 | 0.21 | | 0.31 |
| *Note:* Differences in RSFA across study sample (all genetic status groups combined) following robust multiple linear regression analysis on component-based RSFA maps. Estimated regression parameters, t values, and *p* values are shown for independent components that were not significant after FDR correction at the 0.05 level or were considered as components not related to the predictors of interest in the models, but possibly associated with other factors, such as signals of vascular and CSF origin, and noise signals. Models are adjusted for sex, handedness, and scanning site. | | | | | | | | | | | | | | |

| **Table A.2.** Multiple regression analysis results of independent component (IC) subject loadings from independent component analysis (model*‘RSFA_IC_ ~ 1 + Genetic status*Age + Sex + Handedness + Scanning Site’)* across groups of interest | | | | | | | | | | | | | |
| --- | --- | --- | --- | --- | --- | --- | --- | --- | --- | --- | --- | --- | --- |
| **IC Model**  **Adjusted R^2^ Age Genetic Status Genetic Status*Age** | | | | | | | | | | | | | |
|  |  | **β** | **T** | ***P*** | **FDR-**  **corr. *P*** | **β** | **T** | ***P*** | **FDR-**  **corr. *P*** | **β** | **T** | ***P*** | **FDR-**  **corr. *P*** |
| **IC 4 – Posterior cingulate cortex/precuneus** | | | | | | | | | | | | | |
| Sample | 0.62 | **-0.09** | **-3.59** | **<.001** | **.001** | **-**0.05 | **-**1.77 | .078 | .123 | **-**0.05 | **-**2.00 | .046 | .085 |
| *NC vs SC* |  | **-**0.08 | **-**1.83 | .069 |  |  |  |  |  |  |  |  |  |
| *PSC vs SC* |  | **-0.13** | **-3.18** | **.002** |  |  |  |  |  |  |  |  |  |
| *NC vs PSC* |  | **-0.07** | **-2.63** | **.009** |  |  |  |  |  |  |  |  |  |
| **IC 17 – Posterior parietal association areas** | | | | | | | | | | | | | |
| Sample | 0.54 | **-0.13** | **-4.27** | **<.001** | **<.001** | -0.06 | -1.80 | .072 | .098 | **-0.06** | **-2.17** | **.030** | **.048** |
| *NC vs SC* |  | **-0.16** | **-3.08** | **.002** |  |  |  |  |  | **-0.14** | **-2.18** | **.030** |  |
| *PSC vs SC* |  | **-0.22** | **-4.37** | **<.001** |  |  |  |  |  | **-**0.10 | **-**1.76 | .079 |  |
| *NC vs PSC* |  | **-0.09** | **-3.07** | **.002** |  |  |  |  |  | **-**0.03 | **-**1.13 | .258 |  |
| **IC 21– Right lateral prefrontal cortex** | | | | | | | | | | | | | |
| Sample | 0.44 | **-0.11** | **-3.33** | **<.001** | **0.004** | **-0.10** | **-2.68** | **.008** | **.021** | **-0.07** | **-2.31** | **.021** | **.046** |
| *NC vs SC* |  | **-**0.04 | **-**0.70 | .487 |  | **-0.24** | **-3.67** | **<.001** |  | 0.01 | 0.11 | .915 |  |
| *PSC vs SC* |  | **-**0.06 | **-**1.16 | .247 |  | **-0.22** | **-3.13** | **.002** |  | 0.02 | 0.35 | .726 |  |
| *NC vs PSC* |  | **-**0.06 | **-**1.69 | .092 |  | **-**0.01 | **-**0.33 | .742 |  | **-**0.01 | **-**0.42 | .677 |  |
| **IC 23 – Left lateral prefrontal cortex** | | | | | | | | | | | | | |
| Sample | 0.49 | **-0.22** | **-7.22** | **<.001** | **<.001** | **-**0.06 | **-**1.87 | .062 | .168 | **-0.08** | **-2.52** | **.012** | **.044** |
| *NC vs SC* |  | **-0.13** | **-2.42** | **.016** |  |  |  |  |  | **-**0.01 | **-**0.19 | .846 |  |
| *PSC vs SC* |  | **-0.20** | **-4.08** | **<.001** |  |  |  |  |  | 0.07 | 1.23 | .220 |  |
| *NC vs PSC* |  | **-0.18** | **-5.41** | **<.001** |  |  |  |  |  | **-**0.05 | **-**1.61 | .108 |  |
| *Note:* Differences in RSFA across groups of interest following robust multiple linear regression analysis on component-based RSFA maps. Estimated regression parameters, t values, and *p* values are shown for main effects across the entire sample and post hoc tests between sub-groups of interest where relevant. Outcomes of interest are the RSFA-IC loadings associated with ICA components within GM regions where case-control differences are found. Models are adjusted for sex, handedness, and scanning site. *P* values are FDR-corrected at the 0.05 level in comparisons across the whole sample (all genetic status groups combined). Bold numbers indicate that *p* values are statistically significant. NC, non-carrier; PSC, pre-symptomatic mutation carrier; SC, symptomatic mutation carrier. | | | | | | | | | | | | | |

| **Table A.3.** Multiple regression analysis results following voxel-based region of interest analysis (model*‘RSFA_Voxel_ ~ 1 + Genetic Status*Age + Sex + Handedness + Scanning Site’)* across groups of interest | | | | | | | | | | | | | |
| --- | --- | --- | --- | --- | --- | --- | --- | --- | --- | --- | --- | --- | --- |
| **ROI Model**  **Adjusted R^2^ Age Genetic Status Genetic Status*Age** | | | | | | | | | | | | | |
|  |  | **β** | **T** | ***P*** | **FDR-**  **corr. *P*** | **β** | **T** | ***P*** | **FDR-**  **corr. *P*** | **β** | **T** | ***P*** | **FDR-**  **corr. *P*** |
| **Left middle frontal gyrus** | | | | | | | | | | | | | |
| Sample | 0.23 | **-0.09** | **-2.51** | **.012** | **.034** | **-0.14** | **-3.25** | **.001** | **.005** | **-0.16** | **-4.28** | **<.001** | **<.001** |
| *NC vs SC* |  | **-**0.03 | **-**0.53 | .594 |  | **-0.29** | **-3.91** | **<.001** |  | **-**0.13 | **-**1.73 | .084 |  |
| *PSC vs SC* |  | **-0.15** | **-2.39** | **.017** |  | **-0.22** | **-2.70** | **.007** |  | **-**0.07 | **-**0.98 | .326 |  |
| *NC vs PSC* |  | -0.02 | -0.40 | .691 |  | **-**0.03 | **-**0.68 | .499 |  | -0.07 | -1.63 | .103 |  |
| **Right middle frontal gyrus** | | | | | | | | | | | | | |
| Sample | 0.25 | **-0.13** | **-3.72** | **<.001** | **.002** | **-0.15** | **-3.72** | **<.001** | **.002** | **-0.08** | **-2.25** | **.025** | **.041** |
| *NC vs SC* |  | **-**0.07 | **-**1.18 | .239 |  | **-0.31** | **-4.19** | **<.001** |  | **-**0.01 | -0.11 | .909 |  |
| *PSC vs SC* |  | **-0.14** | **-2.22** | **.027** |  | **-0.23** | **-2.96** | **.003** |  | 0.05 | 0.70 | .483 |  |
| *NC vs PSC* |  | **-0.09** | **-2.40** | **.017** |  | **-**0.06 | **-**1.69 | .093 |  | **-**0.05 | **-**1.23 | .220 |  |
| **Left superior frontal gyrus** | | | | | | | | | | | | | |
| Sample | 0.17 | **-**0.05 | **-**1.37 | .170 | .320 | **-0.19** | **-4.45** | **<.001** | **<.001** | **-**0.08 | **-**2.16 | .031 | .093 |
| *NC vs SC* |  |  |  |  |  | **-0.36** | **-4.72** | **<.001** |  |  |  |  |  |
| *PSC vs SC* |  |  |  |  |  | **-0.30** | **-3.65** | **<.001** |  |  |  |  |  |
| *NC vs PSC* |  |  |  |  |  | -0.06 | -1.62 | .105 |  |  |  |  |  |
| **Right superior frontal gyrus** | | | | | | | | | | | | | |
| Sample | 0.26 | **-0.10** | **-2.79** | **.005** | **.013** | **-0.17** | **-4.09** | **<.001** | **<.001** | **-**0.06 | **-**1.54 | .123 | 0.148 |
| *NC vs SC* |  | -0.05 | -0.77 | .441 |  | **-0.34** | **-4.58** | **<.001** |  |  |  |  |  |
| *PSC vs SC* |  | -0.07 | -1.17 | .241 |  | **-0.24** | **-3.06** | **.002** |  |  |  |  |  |
| *NC vs PSC* |  | -0.07 | -1.69 | .092 |  | -0.06 | -1.56 | .118 |  |  |  |  |  |
| *Note:* Differences in RSFA across groups of interest following robust multiple linear regression analysis in several representative ROIs based on voxel-wise univariate analysis on RSFA maps. Estimated regression parameters, t values, and *p* values are shown for main effects across the entire sample and post hoc tests between sub-groups of interest where relevant. Outcomes of interest are the RSFA-ROI values associated with each ROI where case-control differences are found. Models are adjusted for sex, handedness, and scanning site. *P* values are FDR-corrected at the 0.05 level across the whole sample (all genetic status groups combined). Bold numbers indicate that *p* values are statistically significant. NC, non-carrier; PSC, pre-symptomatic mutation carrier; SC, symptomatic mutation carrier. | | | | | | | | | | | | | |

| **Table A.4.** Anatomical localisation of voxel-based analysis derived clusters where RSFA decreases are observed in comparisons between symptomatic carriers, pre-symptomatic carriers, and non-carriers | | | | | | | |
| --- | --- | --- | --- | --- | --- | --- | --- |
| **Cluster** | **Cluster size** | **Cluster level,**  ***p* FWE- corrected** | **Peak-level,**  ***p* FDR-corrected** | **Peak-level T score** | **MNI Coordinates (mm)** | | |
| **Genetic status effect – SC > NC** | | | | | | |  |
|  |  |  |  |  | **x** | **y** | **z** |
| Left middle frontal gyrus | 10905 | *p*<.001 | *p*<.001 | 7.57 | -48 | 12 | 50 |
| Right middle frontal gyrus |  | *p*<.001 | *p*<.001 | 6.45 | 34 | 2 | 66 |
| Right superior frontal gyrus | 2835 | *p*<.001 | *p*<.001 | 6.66 | 0 | 24 | 42 |
| Right superior temporal gyrus | 2813 | *p*<.001 | *p*<.001 | 5.86 | 50 | 18 | -6 |
| Left posterior cingulate cortex | 1392 | *p*=.010 | *p*<.001 | 4.39 | -6 | -48 | 28 |
| Right posterior cingulate cortex |  | *p*=.010 | *p*=.001 | 4.23 | 0 | -58 | 24 |
| **Genetic status*Age effect – SC > NC** | | | | | | | |
| Left middle frontal gyrus | 1101 | *p*=.004 | *p*=.001 | 5.81 | -48 | 12 | 50 |
| Left superior frontal gyrus |  |  | *p*=.008 | 3.85 | -22 | 20 | 44 |
| Left posterior cingulate cortex/precuneus | 3549 | *p*<.001 | *p*<.001 | 5.50 | -4 | -46 | 24 |
| Right posterior cingulate cortex |  | *p*<.001 | *p*=.001 | 4.93 | 0 | -58 | 24 |
| Left dorsal anterior cingulate cortex |  | *p*<.001 | *p*=.004 | 4.14 | -2 | 12 | 40 |
| Right insula | 1316 | *p*=.002 | *p*=.002 | 4.57 | 34 | 16 | -2 |
| Right caudate nucleus |  | *p*=.002 | *p*=.013 | 3.61 | 12 | 6 | 8 |
| **Genetic status effect – SC > PSC** | | | | | | | |
| Right insula | 12046 | *p*<.001 | *p*<.001 | 6.09 | 36 | 14 | -2 |
| Right pre-central gyrus |  | *p*<.001 | *p*<.001 | 5.37 | 38 | 6 | 20 |
| Right superior temporal gyrus |  | *p*<.001 | *p*<.001 | 4.87 | 52 | 16 | -10 |
| Right dorsal anterior cingulate cortex | 1118 | *p*=.043 | *p*<.001 | 5.66 | 0 | 20 | 40 |
| Right rostral anterior cingulate cortex |  | *p*=.043 | *p*=.016 | 3.16 | 2 | 36 | 24 |
| Left middle frontal gyrus/dorsal prefrontal cortex | 3783 | *p*<.001 | *p*=.001 | 4.70 | -34 | 36 | 48 |
| Right middle frontal gyrus | 1197 | *p*=.035 | *p*=.002 | 4.18 | 40 | 22 | 52 |
| Right middle frontal gyrus/dorsal prefrontal cortex |  | *p*=.035 | *p*=.003 | 3.99 | 42 | 38 | 38 |
| **Genetic status*Age effect – SC > PSC** | | | | | | | |
| Right insula | 2996 | *p*<.001 | *p*=.011 | 4.60 | 36 | 16 | -2 |
| Right superior temporal gyrus |  | *p*<.001 | *p*=.011 | 4.57 | 54 | 16 | -10 |
| Left entorhinal cortex | 1014 | *p*=.008 | *p*=.017 | 4.02 | -34 | 2 | -2 |
| Left inferior frontal gyrus |  | *p*=.008 | *p*=.018 | 3.86 | -28 | 2 | -8 |
| Left thalamus |  | *p*=.008 | *p*=.018 | 3.85 | -18 | -18 | -2 |
| **Genetic status/Genetic status*Age effect – PSC vs NC** | | | | | | | |
| No suprathreshold clusters | | | | | | | |
| *Note:* Clusters where RSFA differences are observed between groups of interest based on voxel-wise univariate analysis on RSFA maps. The primary cluster-forming threshold was set at *p* < 0.05. Clusters are named according to their overlap with the Johns Hopkins University (JHU) atlas. FWE, family-wise error; FDR, false-discovery rate. MNI, Montreal Neurological Institute. NC, non-carrier; PSC, pre-symptomatic mutation carrier; SC, symptomatic mutation carrier. | | | | | | | |

| **Table A.5.** Multiple regression analysis results following independent component analysis and voxel-based region of interest analysis (models*‘RSFA_IC/Voxel_ ~ 1 + Gene Mutation*Age + Sex + Handedness + Scanning Site’*) | | | | |
| --- | --- | --- | --- | --- |
| **Predictor of interest Model Adjusted R^2^ β T FDR-corr. *P*** | | | | |
| ***ICs based on Independent Component Analysis*** | | | | |
| **IC 4 – Posterior cingulate cortex/precuneus** | 0.61 |  |  |  |
| Gene mutation |  | **-**0.01 | **-**0.44 | .826 |
| Gene mutation*Age |  | **-**0.03 | **-**1.15 | .341 |
| **IC 17 – Posterior parietal association areas** | 0.53 |  |  |  |
| Gene mutation |  | 0.06 | 1.76 | .112 |
| Gene mutation*Age |  | **-**0.002 | **-**0.07 | .943 |
| **IC 21 – Right lateral prefrontal cortex** | 0.42 |  |  |  |
| Gene mutation |  | 0.02 | 0.46 | .811 |
| Gene mutation*Age |  | 0.002 | 0.06 | .951 |
| **IC 23 – Left lateral prefrontal cortex** | 0.47 |  |  |  |
| Gene mutation |  | **-**0.05 | **-**1.52 | .276 |
| Gene mutation*Age |  | 0.03 | 1.13 | .488 |
| ***ROIs based on Voxel-wise Analysis*** | | | | |
| **Left middle frontal gyrus** | 0.17 |  |  |  |
| Gene mutation |  | 0.02 | 0.38 | .777 |
| Gene mutation*Age |  | 0.02 | 0.62 | .667 |
| **Right middle frontal gyrus** | 0.22 |  |  |  |
| Gene mutation |  | 0.06 | 1.50 | .162 |
| Gene mutation*Age |  | 0.09 | 2.50 | .023 |
| **Left superior frontal gyrus** | 0.11 |  |  |  |
| Gene mutation |  | 0.01 | 0.29 | .797 |
| Gene mutation*Age |  | 0.03 | 0.76 | .665 |
| **Right superior frontal gyrus** | 0.23 |  |  |  |
| Gene mutation |  | 0.02 | 0.51 | .641 |
| Gene mutation*Age |  | 0.02 | 0.58 | .622 |
| *Note:* Differences in RSFA across groups of interest following robust multiple linear regression analysis on component-based and voxel-wise univariate RSFA maps. Estimated regression parameters, t values, and *p* values are shown for main effects across the entire sample for predictors of interest (gene mutation, age, gene mutation x age interaction) and are adjusted for sex, handedness, and scanning site. Outcomes of interest are the RSFA-IC loadings associated with ICA components within GM regions and RSFA-ROI values associated with voxel-based ROIs where case-control differences are found. *P* values are FDR-corrected at the 0.05 level across the whole sample (all gene mutation groups combined). Gene mutations refer to *C9orf72*, chromosome 9 open reading frame 72; *GRN*, progranulin; *MAPT*, microtubule-associated protein tau, in the study sample. | | | | |

| **Table A.6.** Total variance explained and coefficients of each principal component (PC) following Principal Component Analysis on nine cognitive variables | | | | | | | | | |
| --- | --- | --- | --- | --- | --- | --- | --- | --- | --- |
|  | ***PC 1*** | ***PC 2*** | ***PC 3*** | ***PC 4*** | ***PC 5*** | ***PC 6*** | ***PC 7*** | ***PC 8*** | ***PC 9*** |
| Variance explained (%) | **62.32** | 9.12 | 7.00 | 5.06 | 4.45 | 3.92 | 3.36 | 2.84 | 1.94 |
| **Coefficients per Cognitive Measure** | | | | | | | | | |
| *Digit Span Backwards* | **0.31** | 0.53 | 0.06 | -0.05 | -0.10 | 0.74 | 0.01 | -0.21 | -0.07 |
| *Digit Span Forwards* | **0.28** | 0.68 | 0.17 | 0.23 | 0.21 | -0.54 | 0.13 | 0.16 | -0.05 |
| *Trail Making Test Part A* | **0.34** | -0.16 | -0.37 | -0.26 | 0.56 | -0.11 | 0.08 | -0.45 | -0.35 |
| *Trail Making Test Part B* | **0.38** | -0.06 | -0.21 | -0.09 | 0.20 | 0.03 | -0.05 | 0.09 | 0.87 |
| *Digit Symbol Task* | **0.36** | -0.14 | -0.25 | -0.15 | -0.05 | 0.14 | 0.11 | 0.79 | -0.31 |
| *Boston Naming Test* | **0.30** | -0.38 | 0.42 | 0.65 | 0.31 | 0.20 | -0.14 | 0.03 | -0.08 |
| *Verbal Fluency (animals)* | **0.35** | -0.23 | 0.24 | -0.05 | -0.42 | -0.12 | 0.73 | -0.22 | 0.04 |
| *Verbal Fluency (combined)* | **0.32** | -0.09 | 0.53 | -0.54 | -0.14 | -0.18 | -0.51 | -0.03 | -0.06 |
| *Block Design Task* | **0.33** | -0.02 | -0.46 | 0.36 | -0.55 | -0.20 | -0.40 | -0.21 | -0.09 |
| *Note:* Information about nine cognitive variables used to derive a compact measure of cognitive function using PCA and respective coefficients of each principal component. PC 1 is found to explain the largest proportion of shared variance across the nine measures of cognitive performance. | | | | | | | | | |

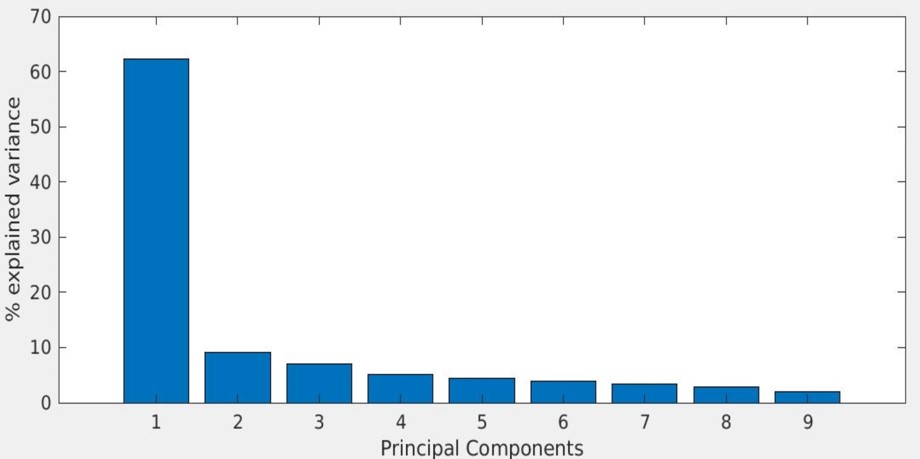

***Figure A.2.*** *Bar graph depicting total variance explained (%) by each principal component following PCA on nine cognitive variables.*

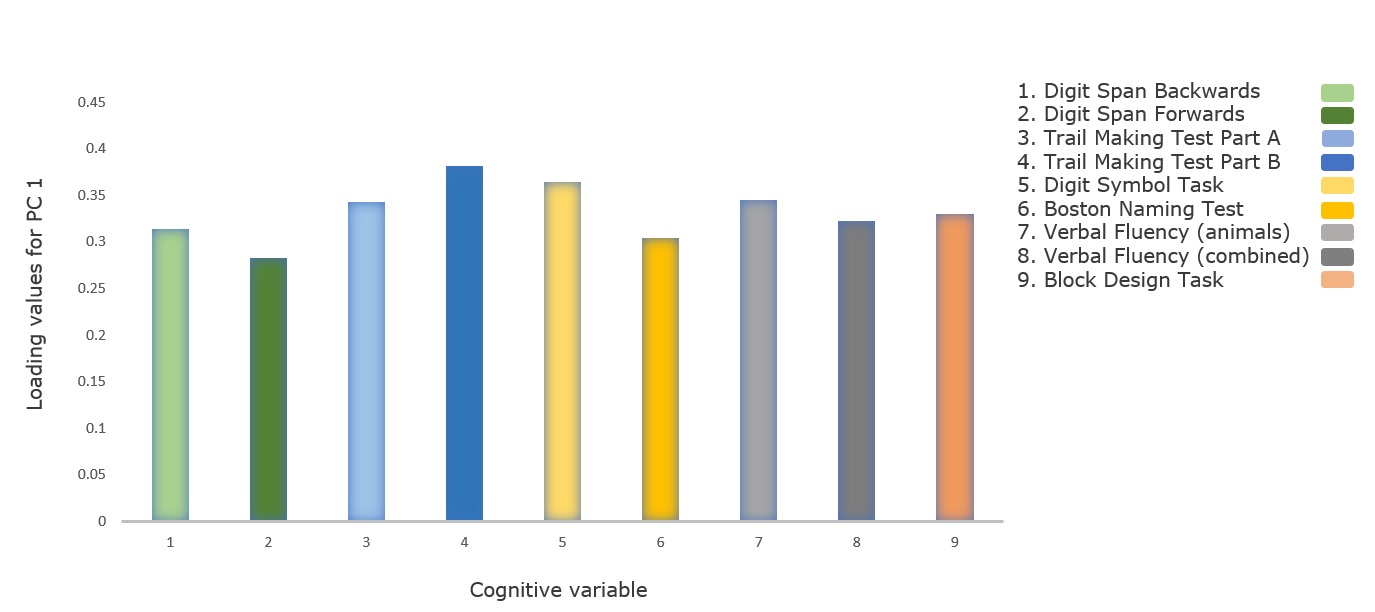

***Figure A.3.*** *Bar graph illustrating subject loadings for the first principal component (PC 1) following PCA, corresponding to nine measures of cognitive performance from the Uniform Data Set, commonly used in neuropsychological assessments in FTD work-up.*
